# An agent-based modelling framework for assessing SARS-CoV-2 indoor airborne transmission risk

**DOI:** 10.1101/2022.07.28.22278138

**Authors:** Simon Knapp, Robert Dunne, Bruce Tabor, Roslyn I. Hickson, Simon Dunstall

## Abstract

We develop a framework for modelling the risk of infection from airborne Severe Acute Respiratory Syndrome Coronavirus 2 (SARS-CoV-2) in well-mixed environments in the presence of interventions designed to reduce infection risk. Our framework allows development of models that are highly tailored to the specifics of complex indoor environments, including layout, people movements, and ventilation. We explore its utility through case studies, two of which are based on actual sites.

Our results reflect previously quantified benefits of masks and vaccinations. We also produce quantitative estimates of the effects of air filters, and reduced indoor occupancy for which we cannot find quantitative estimates but for which positive benefits have been postulated.

We find that increased airflow reduces risk due to dilution, even if that airflow is via recirculation in a large space. Our case studies have identified interventions which seem to generalise, and others which seem to be dependent on site-specific factors, such as occupant density.

## 1. Introduction

Since the start of the Corona Virus Disease of 2019 (COVID-19) pandemic in late 2019, many interventions have been employed to limit the spread of SARS-CoV-2, including vaccinations, masks, physical distancing, physical isolation of contacts, and lock-downs. Numerous simulation studies have been conducted to investigate different aspects of the potential and realised effects of these interventions on a range of outcomes, including disease spread, economic impacts, health, and other social costs. These studies have been conducted at varying spatial scales, from individual rooms and aircraft cabins using Computational Fluid Dynamics (CFD) simulations, through to global scale models developed early in the pandemic to predict rates and patterns of international and intercontinental spread.

Many of these efforts have been focused on population level impacts, for cities [31,34] and nations [7]. Here we present an Agent Based Model (ABM) framework for assessing the risk of SARS-CoV-2 spread via aerosols in indoor environments. Case studies have been chosen to reflect a range of environments, people movements, and potential interventions. Outside of the ABM and cellular automata (CA) literature, significant work has been done on the risk of spread and interventions to mitigate it in indoor environments. Bazant and Bush develop a guideline on the number of people and length of time they may dwell together in a well-mixed space [3]. They use their underlying mathematical framework to estimate quanta emission rates in various super- spreading events of various durations in quite different and complex environments, providing support for and demonstrating the robustness of the underlying assumptions, many of which we rely on here. We build on that work and add the ability to reflect more complex indoor environments, their Heating, Ventilation, and Air Conditioning (HVAC) systems, and the ways individuals interact within them. Rather than producing guidelines aimed at bounding the risk of spread, we produce various metrics on the scale and timing of SARS-CoV-2 spread in specific environments in the presence of a user defined set of interventions.

Given our focus on indoor environments, we discuss existing ABMs and CAs models, and in particular those based on the Wells-Riley model [30, Equation 3]. This implicitly assumes airborne transmission is dominant, a view which was gaining acceptance at the time of writing [11,13,17, 33,42]. We model the airborne viral concentration in each room of an environment through time and calculate the associated risk of infection using a temporally resolved version of the Wells-Riley model, which takes account of the changing viral concentrations susceptible individuals are exposed to. Li et al. [22] have incorporated a spatio-temporal Wells-Riley model into a CFD simulation using the airflow in and layout of a small hospital ward, based on the transmissibility of influenza. They conclude that Respiratory Personal Protective Equipment (PPE) can provide very good protection when the viral load is low and the variant is not very contagious.

A number of ABMs have been targeted at indoor environments. The models with the simplest realistic structure are typified by Cuevas [9] who model agents randomly moving in a 2-dimensional space, with infection risk based on a fixed probability within a radius of infectious individuals. Several models have explored university campuses, with varying levels of detail [2,27,40]. Vecherin et al. develop micro-exposure models of workplaces [36]. Several ABMs consider different types of rooms and buildings [2,36]. Some build schedules based on realistic features such as day of the week, known course schedule (studying or teaching), and residence on or off campus [2], whilst others use surveys to capture social networks [40]. Some models capture the risk of infection using susceptible-infectious-removed progression approaches [27], some use the transition between states as a lognormal distribution [40], whilst others try to capture the epidemiology and physics of interaction [3], and in particular, how close the contact is and for how long [2,36]. The interventions considered by these ABMs range from contact tracing [27], mobility restrictions [9], changes to infection probabilities [9], and combinations of vaccination and other interventions [36].

CA models have been used to explore SARS-CoV-2 spread in indoor settings, though often with different transmission mechanisms and/or interventions than those explored here. Several models use pedestrian dynamics to capture movement within spaces [21,39]. Similarly to the ABMs, the infection processes are based on Wells-Riley approaches [14], capturing multiple modes of transmission, including “direct” and aerosols [21], or progression through states [5,12]. The interventions explored by these CAs models include increased ventilation [21], controlling the numbers or rate of flows of people entering an area [5,21,39], closing indoor eating areas [21], vaccination [12], masks [14], and combinations of non-pharmaceutical interventions [21].

As far as we can ascertain, modelling efforts to date have ignored differences in the way different types of individuals — for example, office staff, receptionists, and visitors — may use and behave in multi-room indoor spaces. In our ABM, individuals inhabit and move through rooms in a manner consistent with their role. At the start of each simulation, one or more individuals are infected and emit virus particles at an activity-dependent rate through time. The virus they emit is instantly mixed into the room’s air and then redistributed through HVAC system(s) and doorways into other rooms and viral concentrations are calculated via coupled differential equations. Non-infected individuals have a dose-dependent probability of infection based on the Wells-Riley Equation. In our office case studies, the simulation stops when a case is detected or the risk of spread becomes insignificant. In contrast, we simulate single days for the museum case study.

Here we present analyses of various interventions designed to reduce the risk of SARS-CoV-2 spread in a hypothetical office and two real-world buildings — another office and a museum. Our framework could be used in any indoor setting and at larger scales if required, provided the context renders the well-mixed room assumption reasonable. Importantly, it allows risk assessment in multi-room environments linked by HVAC systems.

## 2. Methods

Our models are comprised of a set of rooms shared by a group of individuals and which may have one or more HVAC systems and/or air filtration systems installed. On a given day each individual moves between the rooms according to a schedule which is specific to them, may differ each day, and is based on a profile that is specific to their type.

To aid the reader in conceptualising our framework, and to provide a simple example where it is easier to reason about what is happening, we consider a hypothetical office, which is shown in Figure 1. This office is composed of an *open plan* shared workspace, a *meeting room*, a *kitchen*, a *bathroom*, and a *foyer*. We include two types of staff: *office staff* and *receptionists. Office staff* spend most of their time at a desk in the *open plan*, attend meetings in the *meeting room*, occasionally go to the *bathroom*, go to the *kitchen* for lunch once a day, and enter and exit the office through the *foyer* when arriving and leaving work. A *receptionist* is similar, except they spend most of the day in the *foyer* and do not attend meetings. The timing of activity transitions and durations in each room are stochastic and vary between individuals and days. In our hypothetical office we include forty *office staff* and one *receptionist*.

**Figure 1.**
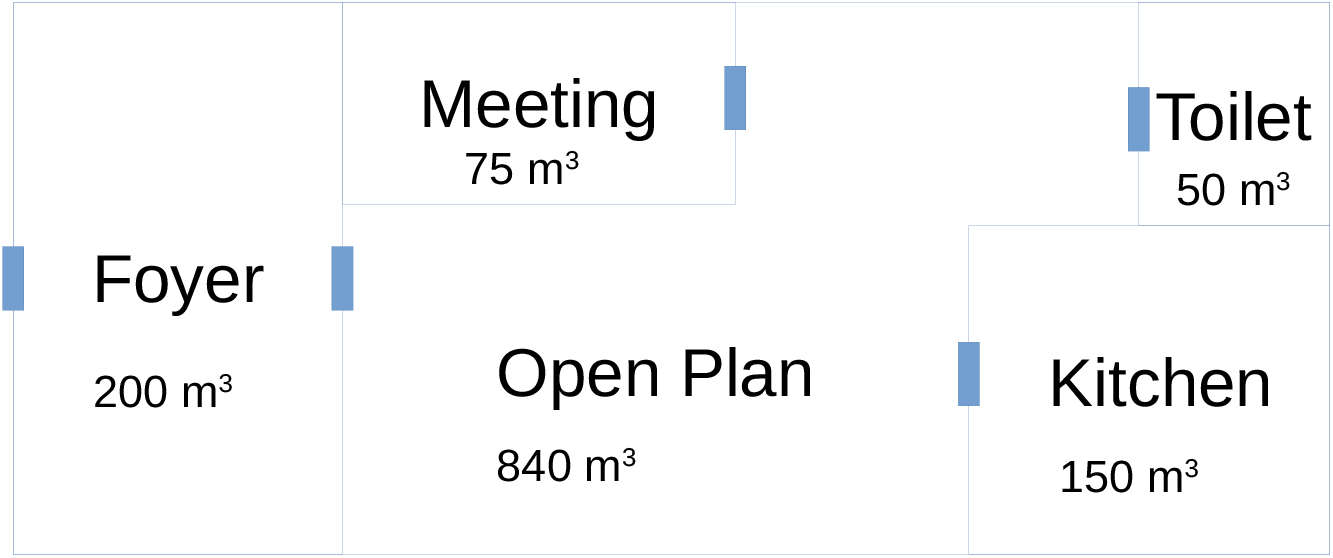
Conceptual diagram of the hypothetical office used in the work presented here (not to scale). Note that the relative positions of the various rooms does not affect this model as we do not consider traversals between boxes.

The framework is designed to make it easy to specify the behaviour of individuals and model the characteristics of different environments. Other types of environments would contain different types of individuals who interact in quite different ways. For example, in a retail store there would be *customer*s and *cashiers*, and on a building site there could be *trades people, foremen, inspectors*, and others. The schedules of these individuals would be comprised of different types of gatherings and activities, and there may be different behaviours affecting air flow and activity levels. The core of the framework simply provides a set of algorithms that execute the following steps (in order) in each day:

i. track the movement of individuals through the rooms of the environment;
ii. track the temporal quanta emissions of each infected individual in each room;
iii. calculate the resulting temporal concentration in each room;
iv. track the exposure of the susceptible individuals;
v. estimate the aggregate infection risk to each susceptible individual; and
vi. randomly infect susceptible individuals based on their infection risk.

## (a) Model Components

### (i) Boxes

The framework generically considers any enclosed space as a *box* and we have two types of boxes: *rooms* and *HVAC systems*. We arbitrarily set the volume of HVAC systems to one cubic meter while many industrial or commercial HVAC units will have an internal capacity much larger than this, the effective air changes per hour (ACPH) within an HVAC unit will be extremely large and this setting will be of little consequence. It would be simple to accurately specify the actual sizes if such information were available.

Boxes have the following attributes

- ***Volume:*** A volume. This is used in the calculation of the airborne viral concentration.
- ***Ventilation Rate:*** The rate air is exchanged between the room and the external environment. Note that this does not include exchange with other boxes and the HVAC system, which is dealt with elsewhere.
- ***Maximum Capacity:*** The maximum number of individuals the room can hold at one time. In most cases this is not specified implying that no upper limit is enforced. The only rooms in which we actively constrain capacity are meeting rooms in the office case studies, and theatres in the museum case study. HVAC boxes have a capacity of zero.

### (ii) Individuals

Individuals have the following attributes

- ***Role:*** A label that describes the role of the individual. The primary use of this label is in identifying the set of individuals who may attend a given type of gathering. For instance, in our hypothetical office, the only type of gatherings we consider are meetings, which may be attended by *office staff*, but not by *receptionists*. We also use an individual’s role to specify some aspects of their behaviour; for example, some individuals will not wear masks in some rooms. While we have not done so here, the role could also be useful for summarising outcomes; for example, which roles infect which others, or the risk of infection to staff in particular roles.
- ***Infected:*** Whether the individual has ever been infected. In the work described here we assume that infected individuals are no longer susceptible. This is reasonable as our simulations are temporally short enough that this assumption is inconsequential.
- ***Time Infected:*** The time the individual gets infected. This is only defined for individuals that have been infected. It is used in conjunction with their incubation period to determine if they have shown symptoms, and viral shedding rates through their infectious period.
- ***Incubation Period:*** The incubation period the individual will experience. This is drawn from a probability distribution specified for the virus at the time the individual becomes infected.
- ***Symptomatic:*** Whether the individual will show symptoms. This is drawn from a probability distribution specified for the virus at the time the individual becomes infected.
- ***Vaccination History:*** The periods in which the individual is vaccinated. Here we only consider the length of time since the last vaccination, but other algorithms could be specified (see Section 3.1 of the supplementary material for how vaccines are modelled).

### (iii) Virus

The virus has the following attributes and the values we have used are based on estimates reported in the literature for the Wild type, the ancestral Wuhan virus that first emerged in 2019 (WT) variant.

- ***Incubation Period:*** A Continuous probability distribution of the incubation period an individual will experience if they become infected. We use a Weibull distribution with shape and scale parameters 3 and 7.2, respectively [1].
- ***Infectiousness Curve:*** Relative infectiousness since time of infection. We use a gamma distribution with shape and scale parameters 3.420 and 1.338 respectively, which were calculated using data from [28]. This is aggregated to days and re-scaled so the daily infectivities sum to one. We use Bazant and Bush [3, supplementary material, Table S1] to convert relative infectiousness to quanta emissions for the WT.
- ***Probability of Symptomatic Expression:*** The probability that an individual will express symptoms if they become infected. Estimates of this quantity vary significantly [e.g. 15], and we use a constant probability of 0.5.

### (iv) Activities

Individuals move between the rooms within an environment based on a schedule that is generated sequentially in the following two steps:

i. ***Gatherings:*** Groups of individuals are selected to spend time in a room together. Face-to-face meetings are an example of a type of gathering. Note that individuals will also spend time together in the same room when they undertake *activities* independently (for example, eating lunch in the kitchen).
ii. ***Activities:*** Individuals choose how to spend the rest of their time in the environment by scheduling activities around the gatherings they are scheduled to attend.

## (b) Disease Transmission

The notation used in this section is described in Table 1.

**Table 1.** Description of the notation for the model components. **9**

| Symbol | Units | Description |
| --- | --- | --- |
| $A$ | $\text{m}^2$ | Area of a room |
| $V$ | $\text{m}^3$ | Volume of a room |
| $C_t$ | $\text{quanta}/\text{m}^3$ | Viral concentration in a room at time $t$ |
| $I_t$ | | The set of infectious individuals in a room at time $t$ |
| $P$ | $\text{quanta}$ | Rate of viral exhalation of each individual as defined in [3] |
| $s_{it}$ | $\text{quanta}$ | Viral exhalation rate of individual $i$ at time $t$ ($i \in I_t$) |
| $q_{it}$ | $\text{m}^3/\text{h}$ | Breathing rate of individual $i$ at time $t$ |
| $Q$ | $\text{m}^3/\text{h}$ | Air outflow rate from a single well-mixed room |
| $Q_r$ | $\text{m}^3/\text{h}$ | Air exchange rate from mechanical ventilation (e.g air-conditioners) |
| $pf$ | | Filtration efficiency of recirculating filters |
| $v_s$ | $\text{quanta}/\text{h}/\text{m}^2$ | Settling speed of virus |
| $\lambda_v$ | $\text{quanta}/\text{h}/\text{m}^3$ | Viral deactivation rate |
| $p_{it}$ | | Probability that individual $i$ becomes infected in a time interval starting at time $t$ |
| $d_{it}$ | $\text{virion}$ | Dose of virus received by individual $i$ in a time interval starting at time $t$ |
| $\mathbf{c}$ | $\text{quanta}/\text{m}^3$ | Vector of airborne viral concentrations in each room |
| $\mathbf{v}$ | $\text{m}^3$ | Vector of room volumes |
| $\mathbf{q}_v$ | $\text{m}^3/\text{h}$ | Vector of air outflow* rates from a set of rooms |
| $\mathbf{q}_r$ | $\text{m}^3/\text{h}$ | Vector of airflow rates through recirculating filters in a set of rooms |
| $\mathbf{Q}$ | $\text{m}^3/\text{h}$ | Matrix of air exchange rates between a set of rooms** |
| $\mathbf{q}$ | $\text{m}^3/\text{h}$ | Vector of air exchange rates from one room into all other rooms in a set of rooms |
| $\mathbf{s}$ | $\text{quanta}/\text{h}$ | Vector of shedding rates in each room |

\*\* To the external environment. \*\* Element $ij$ is the amount of air flowing from room $i$ to room $j$ , diagonal elements are zero.

### (i) Probability of Infection

We model the probability of an individual *i* becoming infected in a particular time interval starting at *t* using the following parameterisation of the Wells-Riley equation

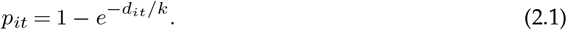

Here, *k* is a disease specific constant that converts virion counts into quanta; that is, this is a scaled dose-response model. We note that *k* and will vary between SARS-CoV-2 variants. We can write *d*^*it*^ as an integral over the rate of exposure, which is proportional to the airborne viral concentration, as

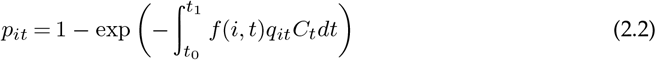

where *t*_0_ is the time the individual enters the environment, *t*_1_ is the time they leave, and *f* (*i, t*) is a function that encapsulates the combined effect of any interventions that affect the amount of virus inhaled by individual *i* at time *t* (for example, by a mask).

### (ii) Airborne Viral Concentration

Bazant and Bush express the rate of change of the airborne viral concentration using the following function of the rate airborne virus is introduced by infected individuals, and the rate it is removed through ventilation, filtration, sedimentation and deactivation [3, Equation 1]:

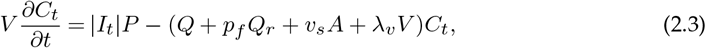

Here we ignore the term *v*^*s*^*A*, representing settling out of the virus particles. This will overstate viral concentrations and hence the apparent efficacy of some interventions at higher airborne viral concentrations. We also replace |*I*^*t*^|*P* with 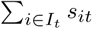 allowing us to include variable shedding rates amongst infected individuals. Hence (2.3) becomes

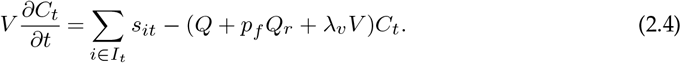

We set *λ*^*v*^ = 0.63 which corresponds to a half-life of 1.1 h. This is the value assumed by Bazant and Bush, based on [35], and is hence consistent with the model based estimates of quanta emission rates we are using here. While the viral deactivation rate is known to vary with temperature and humidity [29] we confine ourselves to a single estimate to reflect the relatively uniform conditions of an air-conditioned indoor space.

Equation (2.4) is specified for a single well-mixed room, but can be naturally extended to a multi-room setting allowing for airflow between the rooms and between the rooms and the HVAC system as

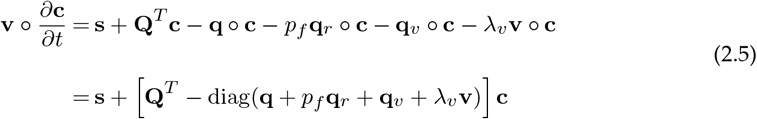

where ○ denotes the Hadamard product.

We can include any number of HVAC systems in a model. In our office case studies, we assume the building is serviced by a single HVAC system. In our museum case study, we include multiple HVAC systems, with detail outlined in Section 4.2 of the supplementary material.

### (c) Case Studies

Our case studies were chosen to reflect a range of environments, people movements and potential interventions. The buildings we explore are the top floor of the Commonwealth Scientific & Industrial Research Organisation (CSIRO) office in Sydney, and Questacon: The National Science and Technology Centre in Canberra. The CSIRO office is a typical office and allows us to evaluate the interventions we explore in our hypothetical office in a real office. The details of the CSIRO office and how we have modelled it are included below and in Section 4.1 of the supplementary material. Questacon receives hundreds to thousands of visitors per day and provides an opportunity to evaluate similar interventions in a very different environment. The physical layout of Questacon also provides opportunities for exploring the challenges associated with describing and modelling a building with complicated HVAC systems and people movements. Brief details of Questacon and how we have modelled it are included below and further detail is provided in Section 4.2 of the supplementary material.

#### (i) CSIRO Office

The CSIRO office is a fairly typical office which occupies the top three floors of a building in Eveleigh, New South Wales (NSW). We have modelled an office based on the layout of the fifth floor, shown in Figure 4 of Appendix A of the supplementary material, and the lunchroom on the fourth floor. The office has a total volume of approximately 5,000 m^3^. There are 82 *office staff*, with 6 open floor plan areas, 39 private offices, and 7 meeting rooms with a total meeting capacity of 55 individuals.

**Figure 4.**
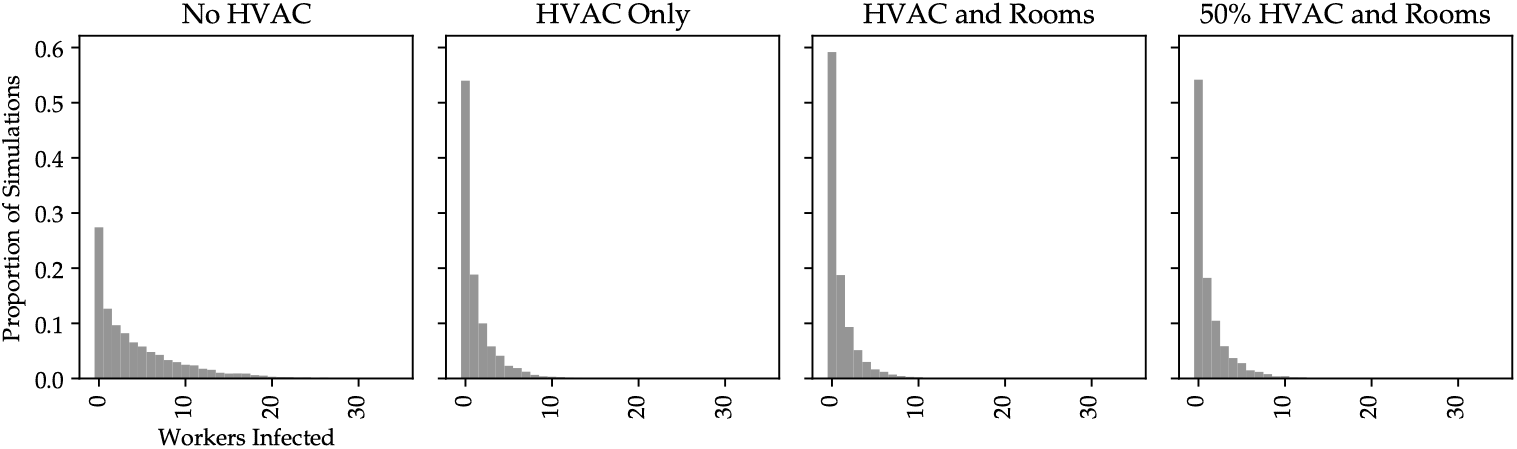
Number of individuals infected in our hypothetical office under different HVAC configurations.

#### (ii) Questacon

Questacon is the National Science and Technology Centre located in Canberra. The primary facility — and the subject of the analyses presented here — is an interactive science museum located in Parkes in the Australian Capital Territory. It is open 9 AM to 5 PM every day of the year except Christmas Day. Daily visitor numbers rival or exceed that of most offices and cumulative annual visits before the pandemic exceeded the population of Canberra. Most visitors will not be regular, and the average visit is around two hours.

During the bushfires in early 2020 dense smoke penetrated the existing F5^1^ filters installed in the Questacon HVAC systems, resulting in an unpleasant staff and visitor experience. Subsequently, F7 filters were installed which are the highest grade that can be fitted to the current HVAC system. These capture 85% of PM2.5 particles (2.5 µm or less), which is the size range of the respiratory aerosols responsible for transmitting SARS-CoV-2.

Gallery staff arrive at 9 AM and leave at 5 PM and remain in a single room for the entire day.

We model the movement of visitors through the building using the following algorithm.

- The number of visitors in a day is specified.
- Given the number of visitors in a day, groups of visitors are generated. Visitor groups are assumed to be of sizes one through five, each with equal probability. Each group starts their journey in the foyer where they dwell for five minutes.
- For each group, the dominant direction of their journey through the galleries is specified based on a draw from a Bernoulli distribution, where 80% of groups start at Gallery 1 (G1) and work their way down to Gallery 8 (G8), and the remainder work their way up from G8 to G1. This split is based on advice from Questacon staff.
- Each time the group enters a gallery, the duration of their stay is drawn from a normal distribution with mean and standard deviation which depends on how many times they have already visited the gallery. On the first visit, they spend ten minutes in the gallery with an standard deviation of three minutes, and on the second visit, they spend three minutes on average with a standard deviation of one minute. Any additional visits are ignored.
- The group then moves to either the previous or next gallery, or to the foyer, depending on which gallery they are in. The probabilities with which they make transitions are shown in Table 2.
- The groups trip through the galleries is finished the first time they arrive in the foyer after visiting Gallery 4 (G4).
- The group then visits the café with probability 70%. If they do, they spend ten minutes in the café (ordering and waiting for their order), then 20 minutes eating in either the foyer or the café with probabilities 55% and 35% respectively (there is a 10% chance they leave the building to eat).
- The trip is finished and they leave the building.

**Table 2.**
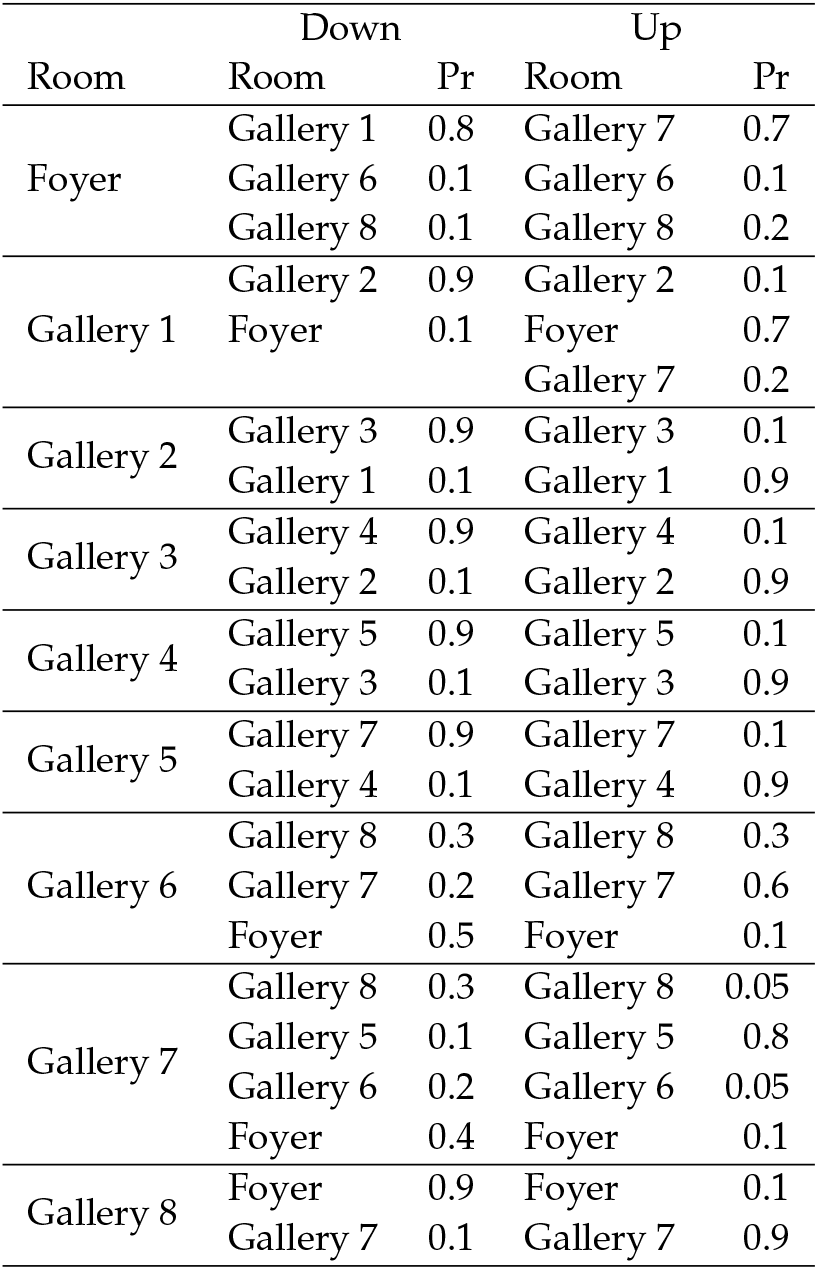
Probabilities of a group transitioning between galleries. The first column is the room they are currently in and the other columns show the probabilities of transitioning to other rooms. The “Down” column corresponds to the case that they start at the top of the ramp around “The Drum” and work their way down, and the “Up” column corresponds to the case that they start at the bottom of the ramp and work their way toward the top.

Presentations are given in the Japan Theatre twice a day at 11 AM and 2 PM. These last for half an hour and any group in the building at the time may attend provided there is enough seating. Those that attend a presentation may spend up to five minutes in the foyer waiting to be let in.

### (d) Interventions

We explore three sets of interventions. The first, *impact of HVAC system configuration and assumptions*, helps us understand the importance of accurately representing the air flows within the environment and hence informs the detail required to develop useful models. The second, *office interventions*, looks at some commonly discussed interventions in the context of an office environment, where a set of individuals share the same environment for many days on end. The third, *museum interventions*, looks at a similar set of interventions in a very different environment with lots of transient visitors.

#### (i) Impact of HVAC System Configuration and Assumptions

The HVAC system installed in a building will have a strong impact on the airborne viral concentrations. We include air exchange directly between rooms, and between rooms and the HVAC system. A challenge in modelling the HVAC system of a building is how one deals with doorways and other connections between rooms. Doors may be partially open or closed, how often and vigorously they are opened and closed will vary, as will the frequency with which individuals traverse doorways. These factors will all affect air-flow, as will changes in mechanically forced or passive ventilation. This detail is hard to generalise and would usually change through the course of a day. We cannot hope to capture the exact state of any environment with respect to these details.

In these analyses we explore the impact of different hypothetical HVAC system configurations and direct air exchange between rooms with the intention of determining the relative importance of the inter-room and mechanically forced flows. We present simulation analyses of airborne viral concentrations and the number of individuals infected. The scenarios we consider are:

- ***No HVAC***: air is exchanged directly between each room and the external environment at a rate of one ACPH (there is no HVAC system).
- ***HVAC Only***: air is exchanged with the external environment at a rate of one ACPH via the HVAC system and air is exchanged between each room and the HVAC system at a rate of seven ACPH. This is in line with the prescription of the Australian Standard AS 1668.2-2012 [32] for the mechanical ventilation of workplaces.
- ***HVAC and Rooms***: air is exchanged with the external environment at a rate of one ACPH via the HVAC system, air is exchanged between each room and the HVAC system at a rate of seven ACPH, and air is exchanged between adjacent rooms via doorways.^2^
- ***50% HVAC and Rooms***: the same as scenario i, except air is exchanged between each room and the HVAC system at a rate of 3.5 ACPH.

#### (ii) Office Interventions

We consider the following interventions, which are commonly discussed in the literature and public debate, in our hypothetical and the CSIRO offices. Each scenario is based on the Business As Usual (BAU) with the modifications noted.

- ***BAU***: This is the ‘no control scenario’. Ventilation is configured as described in the *HVAC and Rooms* scenario described above, infected individuals only get tested if they show symptoms^3^, and meetings can be held in the meeting room.
- ***Masks Only***: All employees wear masks except when they are in the kitchen.^4^
- ***Testing Only***: The entire workforce is tested each day.
- ***Masks and Testing***: All employees wear masks except when they are in the kitchen and are tested each day.
- ***No Meetings***: Meetings are not scheduled; that is, the meeting room is unused.
- ***Vaccination***: All staff are vaccinated at uniformly distributed times over the previous six months.^5^
- ***Increased Ventilation***: Increase the rate at which air is exchanged with the outside environment from one to seven ACPH (which is the rate at which air is exchanged between the rooms and the HVAC system under the BAU).
- ***HVAC Filters***: F7 filters are fitted to the HVAC system outlets, so that air returned from the HVAC systems is filtered with an efficiency of 85% [41].
- ***Portable Filters***: Portable High Efficiency Particulate Air (HEPA) filters with an efficiency of 95% and a capacity of 350 m^3^*/*h [10] are placed in every office, open plan area, meeting room, and kitchen.

#### (iii) Questacon Interventions

We consider the following scenarios, some of which reflect the specifics of Questacon and interventions which have been implemented there. Each scenario is based on the BAU with the modifications noted.

i. ***BAU***: We assume 1,400 visitors a day, 28 of which (corresponding to a 2% community infection rate) were infected four days prior. There are no HVAC filters, all galleries are open, groups that purchase food can eat in the café or foyer, and the Japan Theatre can operate at full capacity (which is 120).
ii. ***Close Mini Q***: Mini Q (Gallery 7 (G7)) is a busy, interactive gallery where children engage in boisterous activities and hence it posed relatively high transmission risk. In this scenario, it is closed.
iii. ***Ban Eating Indoors***: Groups that purchase food in the café always leave the building to eat.
iv. ***Ban Eating Indoors and Close Mini Q***: This is the combination of interventions (ii) and (iii).
v. ***350 Visitors per Day***: Limit the number of visitors to 350 per day and limit the number of available seats in the Japan Theatre to 77.
vi. ***Existing HVAC Filters***: Take account of the existing F7 filters.
vii. ***Existing HVAC Filters, Ban Eating Indoors, and Masks***: This is the combination of interventions (iii) and (vi), and individuals wearing masks at all times. Masks are assumed to reduce both shedding and ingestion by 50% [4].^6^
viii. ***Existing HVAC Filters, Masks, Ban Eating Indoors, and Reduced Visitors***: This is the combination of interventions (v) and (vii) and reflects the set of interventions in place at Questacon at the time of writing.
ix. ***Infected Staff Member***: We reduce the number of infected visitors by one (to 27) and also infect one, randomly selected, staff member. This is not an intervention but an exploration of the relative impacts of different types of individuals becoming infected.

### (e) Simulation Method

We explore interventions through simulation. All simulation experiments are based on 10,000 simulations and we assume a quanta emission rate twice that of WT, which was broadly assumed to be the relative infectiousness of the delta variant [37].

In our office case studies, we present the total number of staff infected and the time until a case is detected as the outcomes of our simulation experiments. These two measures would be of interest to potentially different audiences. The total number of staff infected is likely to be of interest to all decision makers. It is of direct interest to employers and individuals as it is a measure of direct impact on these groups. The time until the first case is detected may be of less interest to these groups, but is likely to be of great interest to public health officials who are also concerned with risks to, and impacts on, the broader community.

In each simulation, we infect a single individual at time zero and run the simulation until either an infected individual is detected, or when no individual has a probability of being infected which was greater than 1/10,000 and all infected individuals have reached the end of their infectious period.

In our museum case study, we simulate single days and report the average number of infections over the simulations within each room and in total.

## 3. Results

### (a) Impact of HVAC System Configuration and Assumptions 19

The impact of various hypothetical HVAC configurations on airborne viral concentrations when four randomly selected staff are infected one through four days before the period shown are shown in Figure 2. The viral shedding of these four staff is shown in Figure 3.

**Figure 2.**
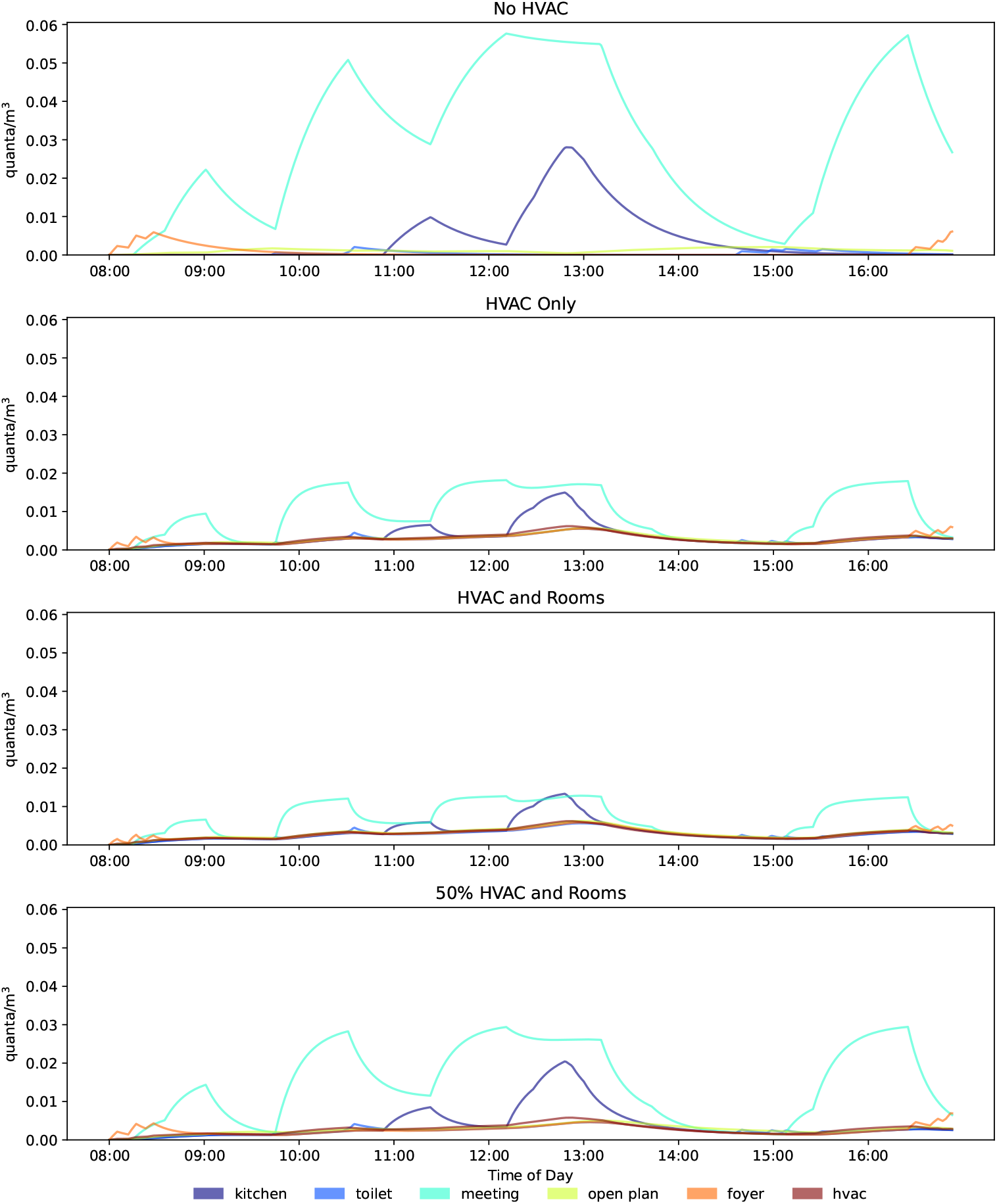
Airborne viral concentrations in the rooms of the hypothetical office under the different HVAC configurations.

**Figure 3.**
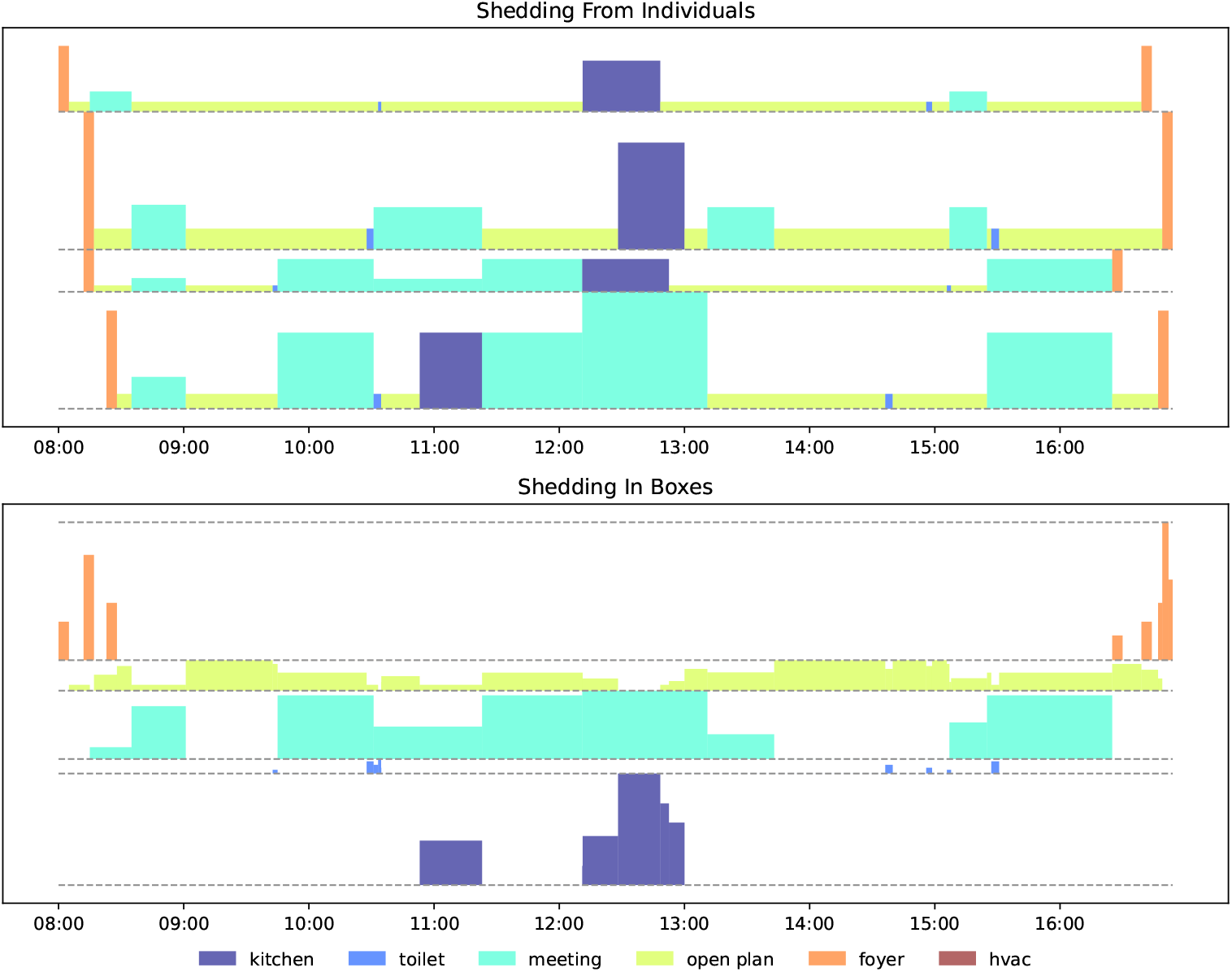
Viral shedding from infected individuals in the hypothetical office for the results shown in Figure 2. Each line in the top panel shows the shedding for an individual through time. The height of the boxes shows the relative amount of viral shedding, and the color denotes the room. The bottom panel shows the aggregated shedding in each room from all individuals.

Comparison of the top panel of Figure 2 to the other panels shows how dramatic the effect of the HVAC system can be, cutting the peak airborne viral concentrations by at least a factor of two and up to a factor of about five. Comparison of the middle two panels shows that the majority of the reduction in concentration is induced by the mechanical ventilation, with the effect of flow through doorways being an almost second order effect.

Figure 4 shows distributions of the number of staff infected under the various scenarios and Table 3 shows the corresponding average number of staff infected and times until an infected staff member was detected. Again, the large impact that airflow has on infection rates is clear.

**Table 3.** Average number of workers infected and average first period in which a case is detected for each HVAC configuration for the hypothetical office. The averages are taken over the simulations used to generate the histograms shown in Figure 4. In the case of the average first period in which a case was detected, these data only include simulations in which an infected worker was detected.

| intervention | Average Day Finished | Average Number of Cases | At Least One Case Detected (%) | Number Infected Reduction From BAU (%) |
| --- | --- | --- | --- | --- |
| No HVAC | 9 | 4.26 | 86 | 0 |
| HVAC Only | 9 | 1.22 | 69 | 71 |
| HVAC and Rooms | 9 | 0.95 | 66 | 78 |
| 50% HVAC and Rooms | 9 | 1.24 | 70 | 71 |

### (b) Office Interventions

Figures 5 and 6 show the distributions of the total number infected and the times of detection for our hypothetical office, for each of the interventions we have explored; Figures 7 and 8 show the corresponding results for the CSIRO office; and Table 4 summarises these data.

**Table 4.**
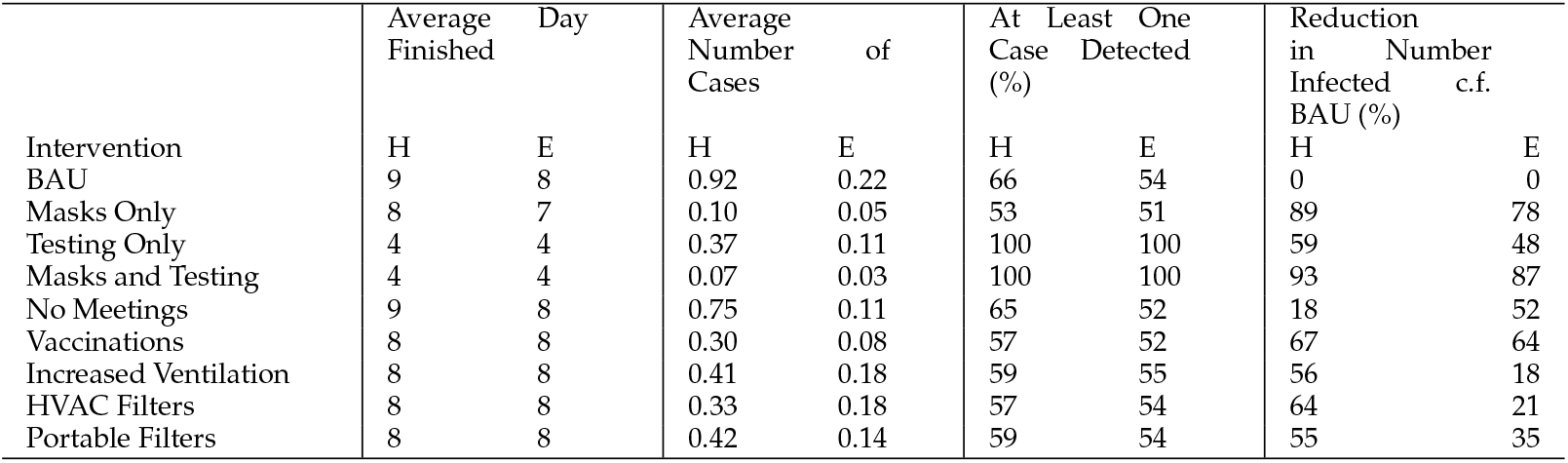
Average number of workers infected and average first period in which a case is detected for each intervention for each site considered. Here “H” refers to the hypothetical office, and “E” to the CSIRO Eveleigh office.

**Figure 5.**
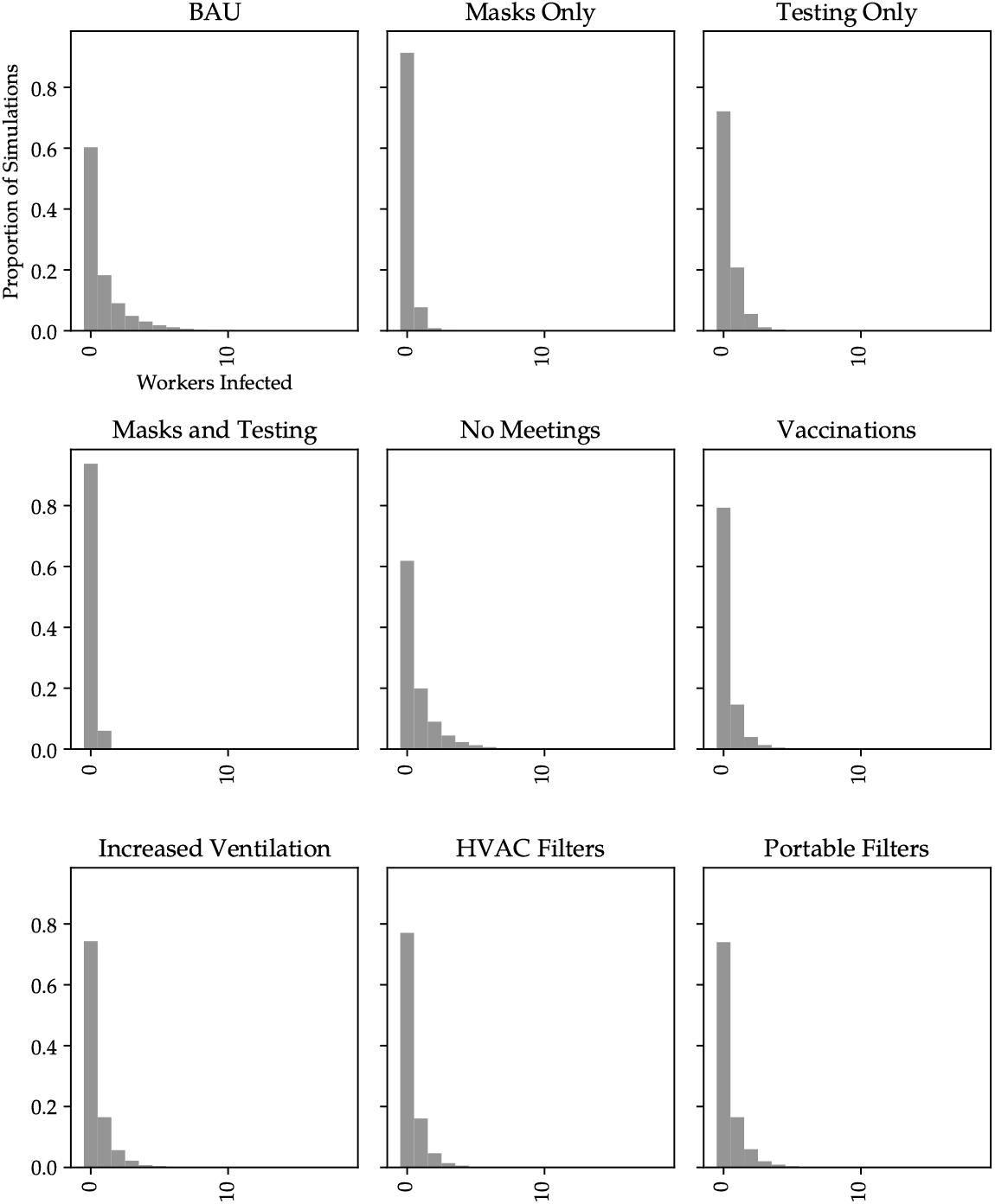
Distributions of the number of staff infected in the various interventions at the time each simulation is terminated for our hypothetical office.

**Figure 6.**
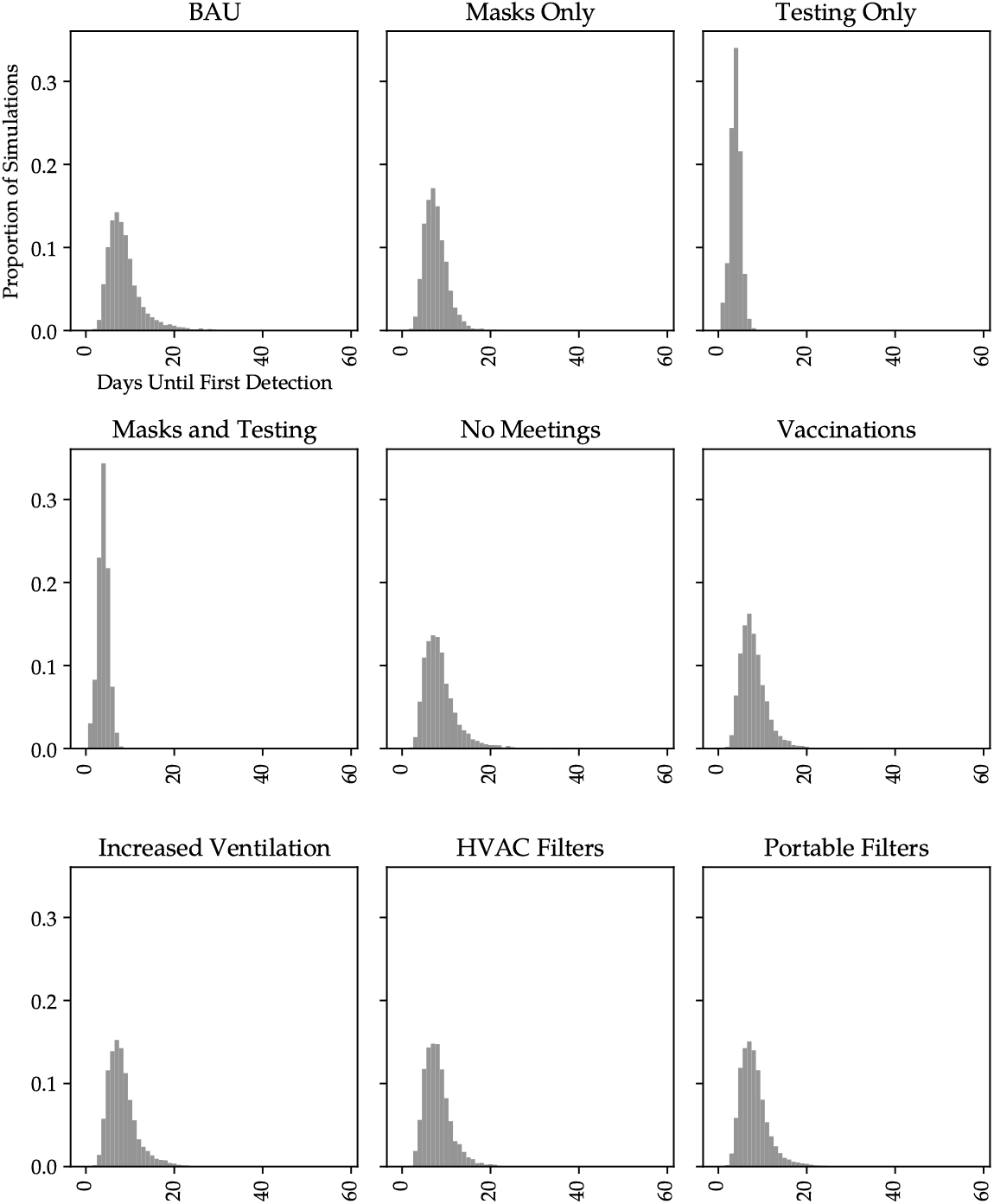
Distributions of the number days until first case detection in the various interventions using for our hypothetical office. Only simulations where at least one case was detected are included.

**Figure 7.**
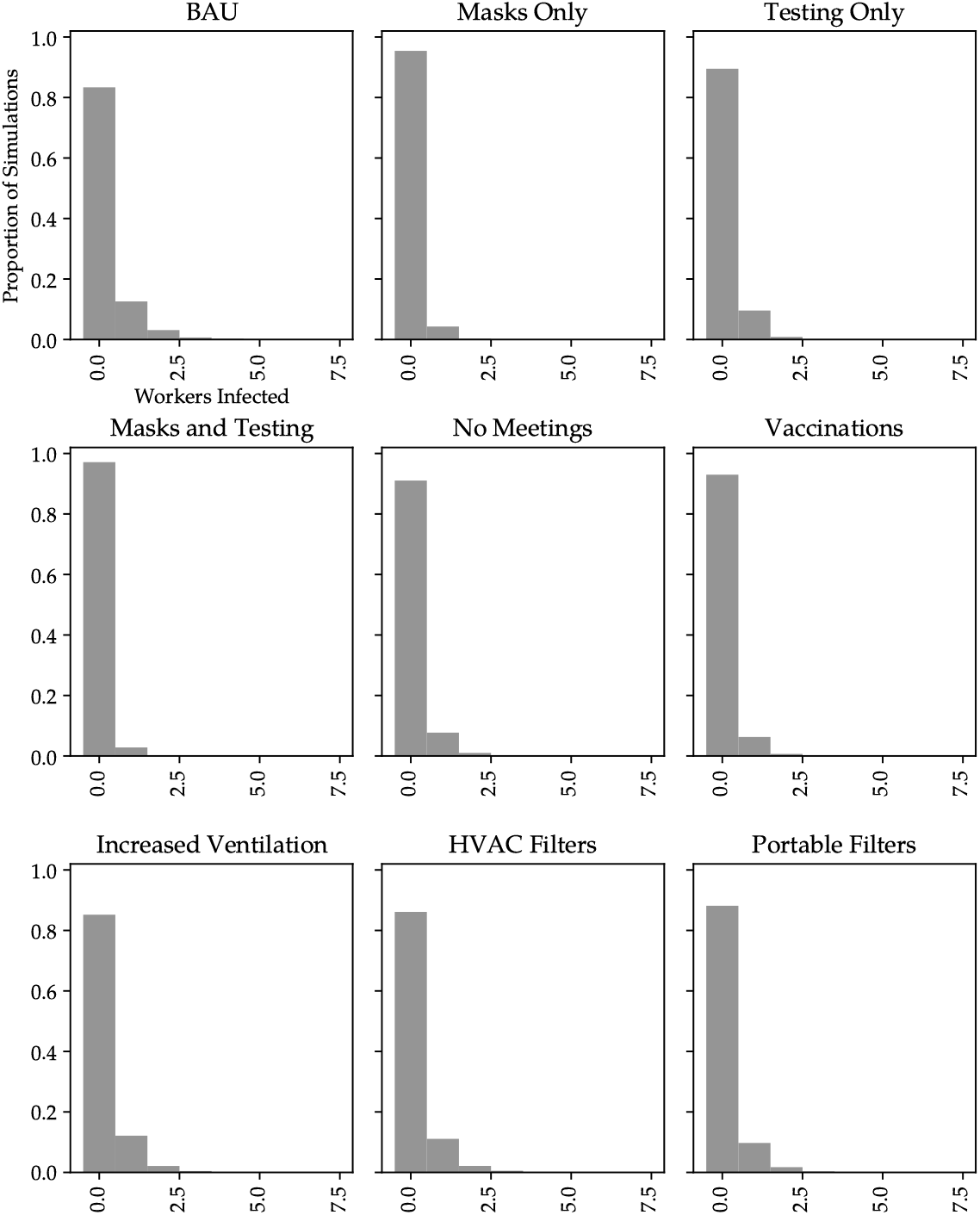
Distributions of the number of staff infected in the various interventions at the time each simulation is terminated for the CSIRO office.

**Figure 8.**
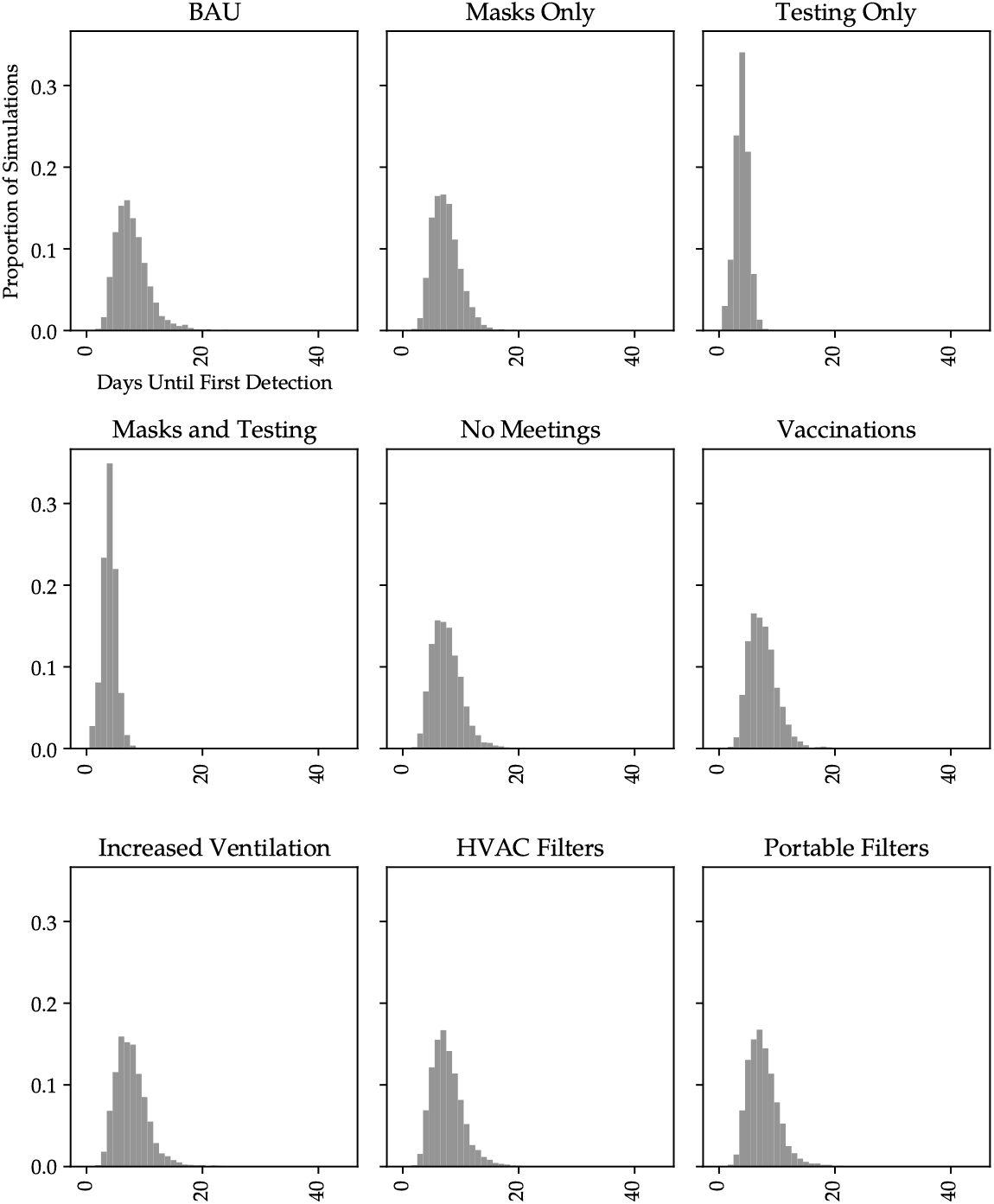
Distributions of the number days until first case detection under the various interventions in the CSIRO office. Only simulations where at least one case was detected are included.

Most interventions significantly reduce the number of infections. Masks, in particular, have a dramatic effect, reducing the number of infections by around an order of magnitude. These results are consistent with both the theory and findings reported elsewhere in the literature. The scenarios including testing (*Testing* and *Masks and Testing*) both approximately halve the time it takes to detect a case and hence, depending on the community response to cases, may be of particular interest from a public health perspective.

Comparison of the columns of Table 4 reveals similar orderings of the impacts of half of the interventions, with notable differences in the others. *No Meetings* is approximately three times as effective in the CSIRO office as it is in our hypothetical office. We believe the main reason for this is that in the hypothetical office the total time spent in meetings is limited due to there being only one meeting room of capacity five. No such restriction exists in the CSIRO office.

The other notable differences are *Increased Ventilation, HVAC Filters* and *Portable Filters*, which are significantly more effective in our hypothetical office than they are in the CSIRO office. The baseline SARS-CoV-2 concentration is lower in the CSIRO office, as the combined volume of all rooms and other spaces is significantly larger relative to the number of staff, and in particular the number of infectious staff (that is, the staff per volume is higher). Consequently, the gains from introducing extra fresh air or increased HVAC efficiency are also lower.

Both the ordering and magnitude of the effects of the interventions on the distributions of the first period in which a case is detected are similar in the two offices.

### (c) Questacon Interventions

Figure 9 shows a summary of a random day at Questacon. The panels show the shedding, viral concentrations and expected number of infections within the various public spaces. The heights of the shaded regions in the top panel are identical because the activity level of all individuals is the same in all locations and they are infected the same length of time prior to the visit. We note that we have observed similar patterns across many simulations. The risk is high in Gallery 5 (G5) because the viral concentration is high, which occurs because it is has a relatively small volume (1,197 m^3^) compared to the other public areas. In contrast, risk in the Foyer is relatively high, but the concentration is not particularly high due to its large volume (5,880 m^3^), and the risk is driven by the amount of time susceptible individuals spend there.

**Figure 9.**
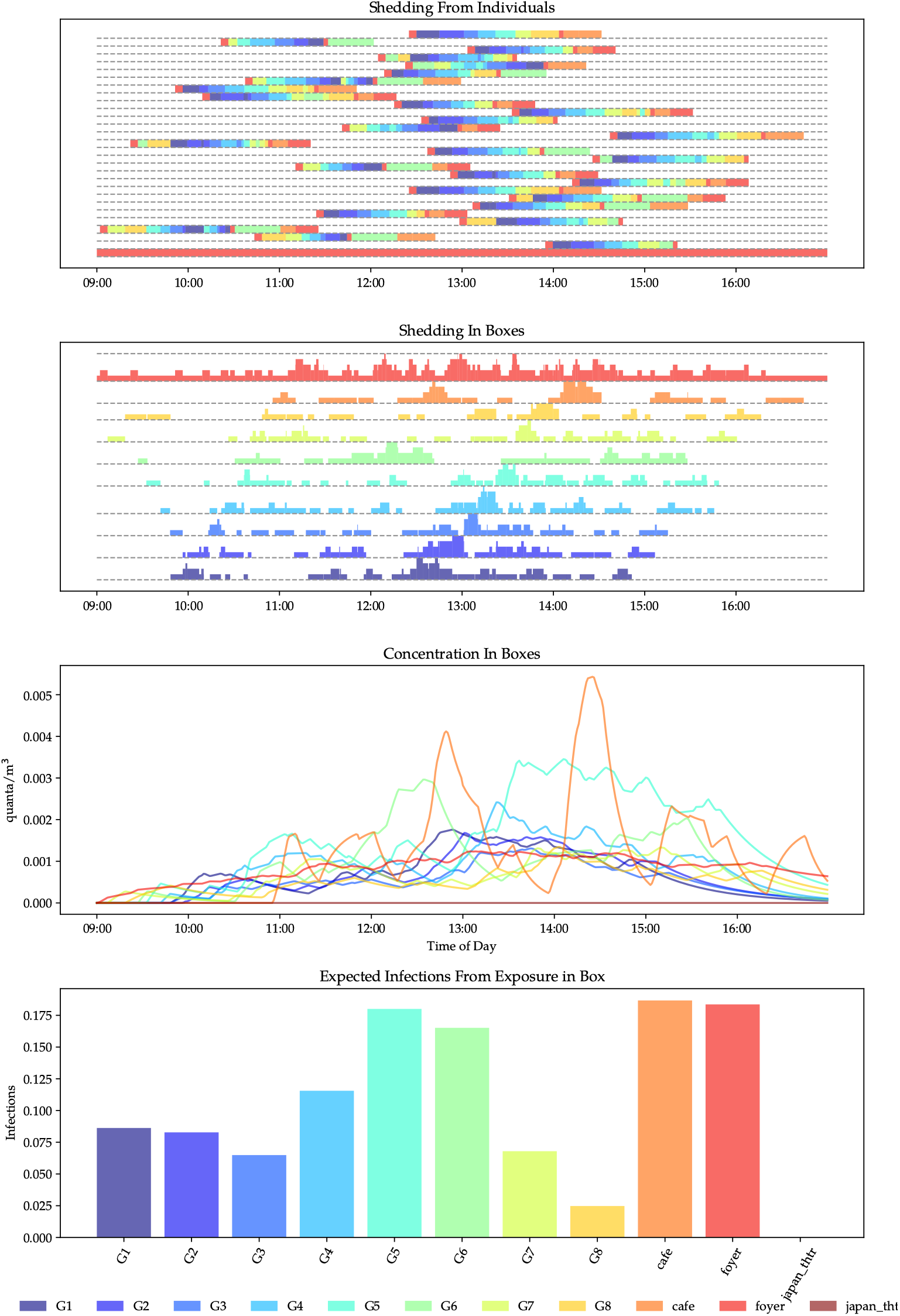
Viral shedding and concentrations in the galleries, theatrettes, café and foyer of Questacon for a simulation of a single day. Top panel: viral shedding from infected individuals. Each line is for a single individual. The color of each shaded region denotes the room they are in and the height shows the relative shedding. Upper middle panel: viral shedding within rooms. This is the aggregated shedding from each infected individual in each box, Lower middle panel: viral concentrations within rooms. Bottom panel: expected number of infections from exposure within each room.

Figure 10 shows box plots of the distributions of the number of infections in each public space for each of the scenarios. Table 5 shows the corresponding mean number of infections. While it is clear that the interventions have a large effect on the relative risk of infection within each room, the absolute risk is not reduced significantly.

**Table 5.** Average daily number of infections in Questacon under the BAU and for each intervention.

| Scenario | Average Number Infected |
| --- | --- |
| BAU | 0.88 |
| Close Mini Q | 0.71 |
| Ban Eating Indoors | 0.73 |
| Ban Eating Indoors and Close Mini Q | 0.55 |
| 350 Visitors per Day | 0.24 |
| Existing HVAC Filters | 0.19 |
| Existing HVAC Filters, Ban Eating Indoors, and Masks | 0.17 |
| Existing HVAC Filters, Masks, Ban Eating Indoors, and Reduced Visitors | 0.04 |
| Infected Staff Member | 1.01 |

**Figure 10.**
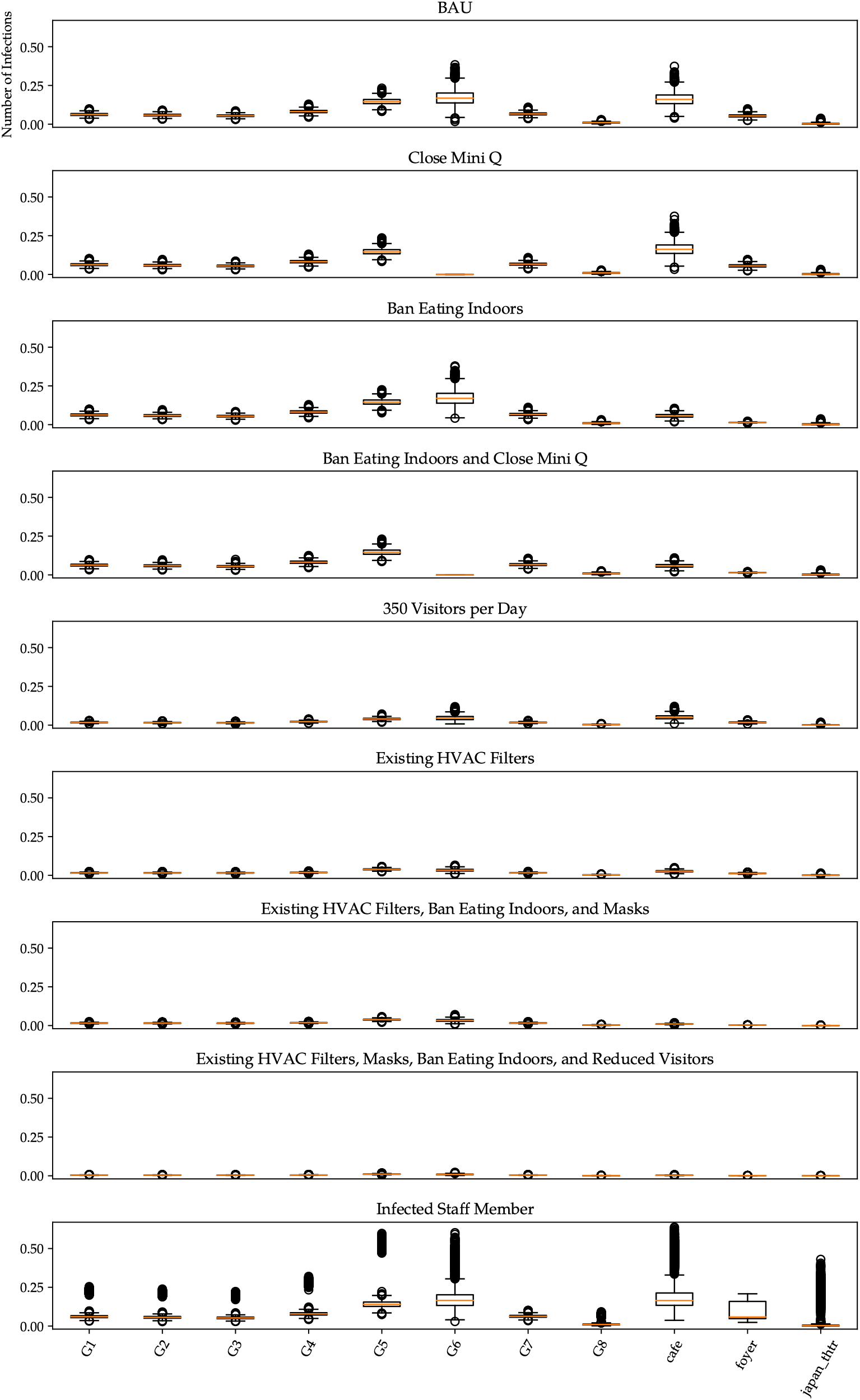
Box plots of the distributions of the expected number of infections due to exposure to airborne virus in each room in Questacon under each scenario.

The risk of infection is significantly higher when there is an infected staff member, most likely due to the long dwelling times in a given location, and the increase in total dwelling time (staff spend three to five times as long in the building as a visitor). This can be seen in the bottom panel of Figure 10, where most of the outliers correspond to simulations where the infected staff member was located in the corresponding room.

## 4 Discussion

We have explored the utility of a new framework for modelling the spread of an airborne virus in indoor settings. We have exercised the framework on a hypothetical office and in two case studies of environments of varying complexity and size. These environments lent themselves to various interventions for minimising the risk of SARS-CoV-2 spread and provided fertile ground for demonstrating and extending the capabilities of the framework.

Our results reflect previously quantified benefits of masks [16] and vaccinations [24]. We also produce quantitative estimates of the effects of air filters [26], and reduced indoor occupancy for which we cannot find quantitative estimates but for which positive benefits have been postulated. We find that increased airflow reduces risk due to dilution, even if that airflow is via recirculation in a large space. Our case studies have identified interventions which seem to generalise, and others which seem to be dependent on site-specific factors, such as occupant density.

Our model yields new and interesting insights into the risk of spread of SARS-CoV-2. It is, however, hard to ascertain the accuracy of our estimates of absolute risk due to the lack of detailed data on real-world events.

An important caveat associated with this work is that we only considered risk of airborne virus in the context of a well-mixed room. Other modes of transmission present additional transmission risks.

We have demonstrated that our framework can be used to model various interventions and produce a range of measures of risk. In the case of our office case studies we infect a single staff member and record how long it takes to detect a case, if a case is detected, and how many staff get infected by the time detection occurs or the risk in the environment becomes negligible. In the case of Questacon, we introduce numerous infectious individuals into the environment and monitor how many infections occur in a single day. We summarise this risk using daily average infection rates and integrated metrics on the risk within each room.

This flexibility in modelling complex environments lends itself to various risk management paradigms appropriate at different phases of a pandemic and to different audiences. For example, the approach of infecting a single staff member is useful when community spread is minimal and there is a strong desire to quickly curtail spread risk, while the approach of introducing infections to mirror infection rates in the broader community may be more useful when spread is more prevalent.

### (a) The Impact of Interventions

The results of our office case studies demonstrate that some interventions may work similarly across many offices and some may depend on site specific features. As shown in Table 4, masks, testing, and vaccination made consistently high reductions in the number of staff infected. Interventions based on reducing viral concentrations in a room (increased ventilation, HVAC filters, and portable filters) appear to be linked to staff density at a site; with increased volume per staff member reducing baseline concentrations and hence the relative impact of those interventions. Similarly, staff schedules and role types may affect the relative performance of interventions, as indicated by the low relative impact of having fewer meetings in the hypothetical office. This provides evidence that offices may be generalised and broad recommendations may be available, conditionally on the density of the office population. Several of the interventions we explored in our office environments showed promise.

Questacon is a large well ventilated building where a relatively large number of individuals spend most of their time moving around. The large volume of the shared spaces and significant movement of air due to mechanical ventilation leads to relatively low viral concentrations and infection risk, even under the BAU. While the interventions we explored here do change the absolute level of risk, reductions in total infections are low especially when the existing F7 filters are taken into account. The risk of infection is increased significantly when there is a single infected staff member. This suggests that regular screening of staff using Point of Cares (POCs) tests may be of value and that, as expected, job roles with higher dwelling times in a specific location pose different risks.

### (b) The Impact of HVAC

There is compelling evidence and a consensus that SARS-CoV-2 can be spread by aerosols [8] and that in some instances mechanical ventilation has directly delivered the virus which infected a person [23]. A significant finding of the current work is the efficacy of HVAC in reducing the spread of SARS-CoV-2 in some environments, even when the rate of air exchange with outside environment is minimal. The main driver for this is the reduction in viral concentrations in more populated shared spaces. While this comes with increased, but relatively low, concentrations in other areas, the nature of the Wells-Riley equation means that the risk of infection is lower overall. This is a novel finding and differs from the arguments of Witts and Coleman [38] and Morawska et al. [25].

In this work we have modelled a statically configured HVAC system and consider filtering of **31** HVAC system return air and using portable filters. However, there are other aspects to consider, including heating/cooling, humidity control, Carbon Dioxide (CO_2_) control, and other dynamic systems that adjust for internal and external conditions.

### (c) Application of the Framework

The Python package and scripts used in this work are available at [18] or from Github [19]. Developing a model for a new environment requires data describing the layout and HVAC systems and implementation of software components to simulate the movements and activities of the individuals who use it. In the current implementation this requires the skills of an intermediate level Python programmer. Depending on how the software is maintained, a library of such components could be developed and shared publicly, gradually reducing the development overhead for additional environments in the future.

Our framework could be used to model any well-mixed indoor space. Potentially interesting examples include: the shops of a high street (shopping precinct), a transport system, an airport terminal, or a school. Indeed, one could model combinations of these simultaneously. This would be an interesting extension to pursue and could inform policy questions like when to close schools or mandate interventions like wearing masks.

### (d) Limitations

The following limitations apply to our results.

#### We are not considering parameter uncertainty

The variation in our results reflects stochastic variation in the movement of individuals and how they respond to the virus. It does not reflect uncertainty in the parameters of the various models we use (for example, quanta emission rates, incubation periods, probability of asymptomatic cases).

#### We do not consider direct contact or short-distance transmission events (plumes, droplets)

Risks from these modes of transmission would be additional to the risk posed by aerosols. This renders our models most applicable when physical distancing is observed and individuals are wearing masks, both of which reduce the risk from these alternative modes of transmission and strengthen the assumption of a well-mixed room.

#### We do not consider fomite transmission

Risk from this mode of transmission will be additional the risks posed by aerosols. Evidence suggests, however, that the risk of transmission through fomites is low [6].

#### We do not consider dynamic aspects of the airflow in buildings

For example, many HVAC systems adjust in response to changing internal and external environmental conditions, and doors are opened and closed throughout the day.

#### We do not consider weekends

This is only relevant for our office case studies; Questacon operates seven days a week and we only consider single days.

#### The sensitivity of POC tests is based on data collated on PCR tests

We use data from a study of Polymerase Chain Reaction (PCR) tests [20] to parameterise the sensitivity of the test. POC tests will generally be less sensitive than PCR tests so our assessments of their impact on spread are probably optimistic.

#### We assume infected individuals are no longer susceptible

This is appropriate when considering shorter time periods as we do here, but would need to be adapted to take into account waning and imperfect immunity in longer-term studies.

#### Model calibration

Although the underlying models used in this work have been fitted by other authors, we have not directly calibrated our models to data. The data required for calibration exists for simpler settings such as single rooms, but more complex spaces are more challenging.

#### Well-mixed room assumption

The Wells-Riley model assumes a well-mixed room. Ideally computational fluid dynamics modelling would be used to better understand when this assumption holds.

## 5 Conclusions

We have demonstrated the utility of our ABM framework for modelling the airborne spread of SARS-CoV-2. This framework is useful for modelling the risk in specific environments and for exploring general principles related to COVID-19 risk management. It allows for the flexible inclusion of different interventions to mitigate risk. As such, our framework could be used to explore specific or archetypal environments.

Our approach brings together data and insights on viral characteristics, breathing rates of people undertaking different activities, vaccine attenuation rates and the specific characteristics of workplaces. The resulting risk analysis may at times differ markedly from our more naive expectations and could guide us in navigate the ongoing risk.

## Data Availability

All data produced are available online at

https://doi.org/10.5281/zenodo.6911600

https://github.com/Sleepingwell/apsrm

## Data Accessibility

The Python package and scripts used in this work are available at [18] or from Github [19]. Additional information are provided in electronic supplementary material.

## Authors’ Contributions. S.K

Conceptualisation, Data curation, Formal Analysis, Investigation, Methodology, Project administration, Software, Supervision, Validation, Visualisation, writing - original draft; R.D.: Data curation, Writing - original draft; B.T.: Data curation, Writing - original draft; R.I.H.: Methodology, Supervision, Writing - review & editing; S.D. Conceptualisation, Funding acquisition, Project administration, Supervision

## Acknowledgements

The authors thank the staff of Questacon and CSIRO building management for providing details of building plans and HVAC systems where available. We’d also like to thank our colleagues for constructive discussions: Dean Paini and Peter Caley, and support staff at Department of Industry, Science, Energy and Resources (DISER).

## Footnotes

1 See Section 5 of the supplementary material for equivalence between filter standards.

2 See Section 1 of the supplementary material for how we model air movement between rooms.

3 See Section 3.3 of the supplementary material for how testing is modelled.

4 See Section 3.2 of the supplementary material for a description of how masks are modelled.

5 See Section 3.1 of the supplementary material for how vaccines are modelled.

6 See Section 3.2 of the supplementary material for a discussion on the effectiveness of masks.

