## Supplementary Material for "An agent-based modelling framework for assessing SARS-CoV-2 indoor airborne transmission risk"

July 2022

#### 1 Inter-room Air Exchange in the Hypothetical and Eveleigh Offices

We calculate the flow between two connected rooms in the following steps.

1. Assuming that a room is well mixed (that is, the Heating, Ventilation, and Air Conditioning (HVAC) system is designed to achieve a well mixed room for thermal comfort).
2. Assume that this well mixed state is attained in a time span that depends on the air-flow creating currents at the scale of the room (about 10 minutes, see Supplementary Information for [4]).
3. Take adjoining open spaces (open plan and corridor say), and assume that these are mixed when 1/2 of the volume of the smaller space is exchanged with the larger space. Combined with assumption 2, this allows us to solve for the rate of exchange between the rooms.

This gives us a reasonable way of modelling the mixing of air between adjoining spaces. Where there is a door we sample the proportion of time it is open (for an office door) or how often it is opened (for a toilet door) from a uniform distribution on a discrete set of proportions. Another approach would be to include the stochastic nature of airflow by introducing a stochastic "ventilation matrix" (parameter $Q$ in Equation 2.5 of the paper). This could vary throughout a period and more accurately reflect the (variable) airflow though the workplace. However, based on our results, in which mechanical airflow dominates the results, we conjecture that this would induce only a minor change in the results.

#### 2 Generating Schedules for Individuals in the Hypothetical Office and Eveleigh

The method for generating schedules for *office staff* used in our hypothetical office and the Commonwealth Scientific & Industrial Research Organisation (CSIRO) office is as follows:

1. Meetings are scheduled (for all *office staff*). The process for generating meetings is:
   (a) Meetings are scheduled back-to-back for each office. The length of each meeting is based on a mixture of normal distributions with (respectively):
      - means of 20, 45, 60 and 90 minutes,
      - standard deviations of 3, 6, 9 and 9 minutes, and
      - mixing weights of 0.2, 0.35, 0.3 and 0.15 (that is, approximately 20% of meetings are drawn from a normal distribution with mean 20 minutes and standard deviation of 3 minutes, etc.).

   (b) The number of meetings each staff member will attend is generated. Each staff member may attend between one and five meetings each day (inclusive) with equal probability. The staff member may attend fewer meetings if there are insufficient spaces in the full set of meetings to satisfy the demand of all staff.

(c) A staff member is chosen at random and scheduled to attend a meeting, which is also chosen at random from the set of meetings which are not fully subscribed and do not overlap with any meeting they are already attending.

(d) Step 1c is repeated while any staff member has not filled the quota specified for them in Step 1b, and there are still meetings which are not fully subscribed. That is, meetings can be under-subscribed, but where possible staff member quotas are filled.

2. Staff enter the workplace at the beginning of the day through the *foyer*, where they spend five minutes, sometime between 8 AM and 9 AM. This is modelled using a normal distribution with a mean of 8:30am and a standard deviation of ten minutes.

3. Once a day each staff member has a lunch break in the *kitchen*. The lunch break is scheduled around the meetings which the staff member is scheduled to attend. The *ideal* start of the lunch break is modelled using a normal distribution with a mean of 12:30pm and a standard deviation of 20 minutes. The *ideal* length of the lunch break is modelled using a normal distribution with a mean of 40 minutes and a standard deviation of 5 minutes. After generating an *ideal* lunch break, this may be moved and/or shortened to avoid clashes with the meetings the staff member is scheduled to attend.

4. A couple of times a day, each staff member uses the *bathroom*. Bathroom breaks are scheduled around meetings and the lunch break. The length of time between arriving at work and the first bathroom break of the day is modelled using a normal distribution with mean of 3.5 hours and a standard deviation of half an hour. The time until the second and later breaks is modelled using a normal distribution with a mean of four hours and a standard deviation of one hour. The length of a bathroom break is modelled as a normal distribution with a mean of three minutes and a standard deviation of 45 seconds.

5. At all other times, staff are seated at their desk in the *open plan*.

6. At the end of the day, staff leave work via the *foyer*, where they spend five minutes, sometime between approximately 4:31pm and 5:21pm. This is modelled using a normal distribution with a mean of 4:51pm and a standard deviation of ten minutes.

Schedules for the receptionist are generated in a similar fashion, except that in step 5 they are seated in *foyer* and they do not attend meetings.

### 3 Discussion on and Implementation of Interventions

#### 3.1 Vaccinations

Vaccinations could have at least three different impacts on the spread of infection.

Firstly, vaccines may affect infectiousness. The issue still seems to be under debate. [10] present evidence that a vaccinated individual sheds less virus than an unvaccinated individual. [7] report that the duration of virus shedding was significantly reduced in vaccinated individuals compared with unvaccinated individuals, but that the relative rates of shedding of partially vaccinated individuals were more variable. [18] note that the clinical trials of the Moderna and Pfizer vaccines do not provide enough evidence to determine if vaccines reduce infectiousness. It is also possible that the degree to which shedding is reduced varies between strains. Here we have chosen the conservative assumption that vaccinated individuals are equally as infectious as unvaccinated individuals.

Secondly, vaccines may increase the likelihood of asymptomatic cases. [18] note that *"a vaccine with high efficacy against Corona Virus Disease of 2019 (COVID-19) disease but low efficacy against Severe Acute Respiratory Syndrome Coronavirus 2 (SARS-CoV-2) infection, would predominantly convert symptomatic infections to asymptomatic infections."*. They also note that at the time of their writing the efficacy of the Pfizer and Moderna vaccines against SARS-CoV-2 infection was not known.[1] [5] notes that "a growing

[1] Though their efficacy against COVID-19 disease is, of course, better known, depending on the strain, since it is one of the key target quantities in clinical trials.

body of evidence suggests that COVID-19 vaccines also reduce asymptomatic infection and transmission.". Here we assume the probability of asymptomatic cases is not affected by vaccination, which, along with our assumption that vaccines do not affect infectiousness, is equivalent to assuming that the vaccines are equally effective against SARS-CoV-2 infection and COVID-19 disease. If this assumption is not true, we will understate infection risk because testing and hence detection are triggered by the expression of symptoms and asymptomatic individuals will generally not be detected (except in scenarios which include testing of asymptomatic staff) and will have longer to spread the disease.

Thirdly, vaccines reduce susceptibility to COVID-19 disease [12, 11] (though as noted above, perhaps not to SARS-CoV-2 infection). Various mechanisms by which this could occur have been postulated, some of which are:

- Some proportion of vaccinated individuals gain sterilising immunity;
- Vaccinated individuals are able to repel some proportion of exposures;
- Vaccination increases the infectious dose.

Each of these mechanisms would be implemented in a different way and may make a difference to the results, but it is not clear which are at work. Here we assume that the vaccine proportionally reduces the susceptibility of individuals by an amount that varies with time since the last vaccination as shown in Figure 1, which is based on [8].

At the time of writing most individuals who are going to be vaccinated have had the initial two doses and many (if not most) will have also had at least one booster. Hence we ignore the partial effects of having only one of the initial two doses and assume that boosters simply reset the time of the last vaccination.

The efficacy of vaccination also varies between strains. For example, vaccines at the time of writing offer little immunity to Omicron [for example 1].[2]

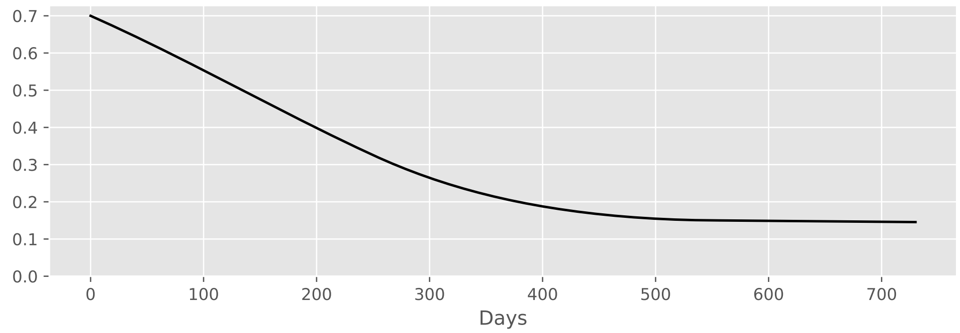


Figure 1: Proportional reduction in susceptibility induced by vaccination as time since last vaccination. The data is based on a derived relationship between the concentration of neutralizing serum antibodies and vaccine effectiveness. For different vaccines and viral strains, the initial effectiveness will vary. See [8].

#### 3.2 Masks

We model masks as providing protection to the wearer of the mask (inhalation efficiency) and also reduction of exhaled pathogen from infected individuals (exhalation efficiency). We introduce some flexibility to reflect the reality that people do not wear masks at all times. In the office scenarios of the current work people wear masks at all times except when they are in a kitchen.

Commonly available masks are: cloth masks, surgical masks, and higher quality masks such as KN95, N95 or P2. There are a wide range of estimates on the performance of each type of mask. The inhalation and exhalation efficiencies of a specific mask may differ as aerosol particles are larger at exhalation and shrink by evaporation due to time spent in the air. Filtration efficiency of smaller particles is poorer in

[2]Vaccines do appear to offer protection against serious illness for Omicron [for example 6], but we are not considering health impacts here.

all but the highest quality masks. Further, some people wear masks which are intended to protect the wearer from things like smoke or dust, which have a valve to ease exhalation and hence exhaled air is not filtered.

Bazant and Bush [4] estimates an efficiency of 30% for cloth masks and 50% for surgical masks. N95 masks nominally exclude 95% of particles of 300 nm. These are worn by hospital staff treating COVID-19 patients. To achieve these performances they must be fit tested, worn properly and be in good condition. We assume variations in design and poorer fitting result in an efficiency of 80%. This is consistent with a recent weekly report from the US Center of Disease Control (CDC) [3], which estimates an efficiency of 83% when such masks are worn by members of the public.

The reported efficiency of masks assumes that mask is in good condition and worn properly. In practice, many people are wearing masks for extended periods of time over multiple days, and often improperly (we have observed, for instance, people wearing the mask over their mouth but not their nose).

In our hypothetical office and the CSIRO office we have assumed that employees will wear a high quality masks and achieve an efficiency of 80% for both inhalation and exhalation. Here we are assuming that staff in these environments can be encouraged to wear high quality masks and will wear them properly. In Questacon we assume an efficiency of 50% for both inhalation and exhalation. This lower value is designed to reflect a broader range of mask types and some level of non-compliance (that is, people not wearing masks or not wearing them properly).

##### 3.3 Testing

The test used for both 'regular testing' (that is, testing that is done when an individual shows symptoms) and systematic testing (for example, testing all staff) is modelled on the results from a study that looked at the sensitivity of a range of Polymerase Chain Reaction (PCR) tests [9]. We model the daily sensitivity of the tests using a Beta-Bernoulli distribution.[3] The data and fitted Beta distributions are shown in Figure 2. In summary, test sensitivity is particularly low on days 1–3 post-infection, before increasing rapidly to peak at around days 7–9 before steadily declining (Figure 3) out until day 21.

In our implementation, individuals implicitly get test results back immediately, which does not reflect the reality of PCR tests. In practice, Point of Care (POC) tests (which individuals would get results back from immediately) are less sensitive, but data on their temporal performance is hard to find.

#### 4 Case Studies

##### 4.1 Eveleigh

CSIRO occupies the top three floors of an office building in Eveleigh, New South Wales (NSW). The building was built in 2008 as the National ICT Australia Ltd (NICTA) building and also houses Defence Science and Technology Organisation (DSTO) on the first two floors. This was chosen as a case study because it is a fairly typical office and we have access to information about it.

We have simulated an office based on the layout of fifth floor of the building, shown in Figure 4 of Appendix A, and the lunchroom on the fourth floor, which is approximately 534 $m^3$ in volume. The fifth floor is serviced by four HVAC units and the kitchen is serviced by one of four HVAC units that service the fourth floor, but we were unable to obtain diagrams describing which rooms are connected to which units and we model the entire office space as serviced by a single, large, HVAC unit.

The Business As Usual (BAU) total occupancy of 170 people over the three floors with no regulation of how many people use the kitchen at one time. The number in meeting rooms is limited only by the capacity of the room. In the work here, we have 82 people in the office, which is based on 70% of the available desks on the fifth floor. We note that, by coincidence, this is twice that of our hypothetical office.

The HVAC system has been modelled as providing seven Air Changes Per Hour (ACPH) of recycled air

[3]Under this model, the probability of a test returning positive given the individual is infected is Bernoulli with parameter $p$, and $p$ is modelled by a Beta distribution.

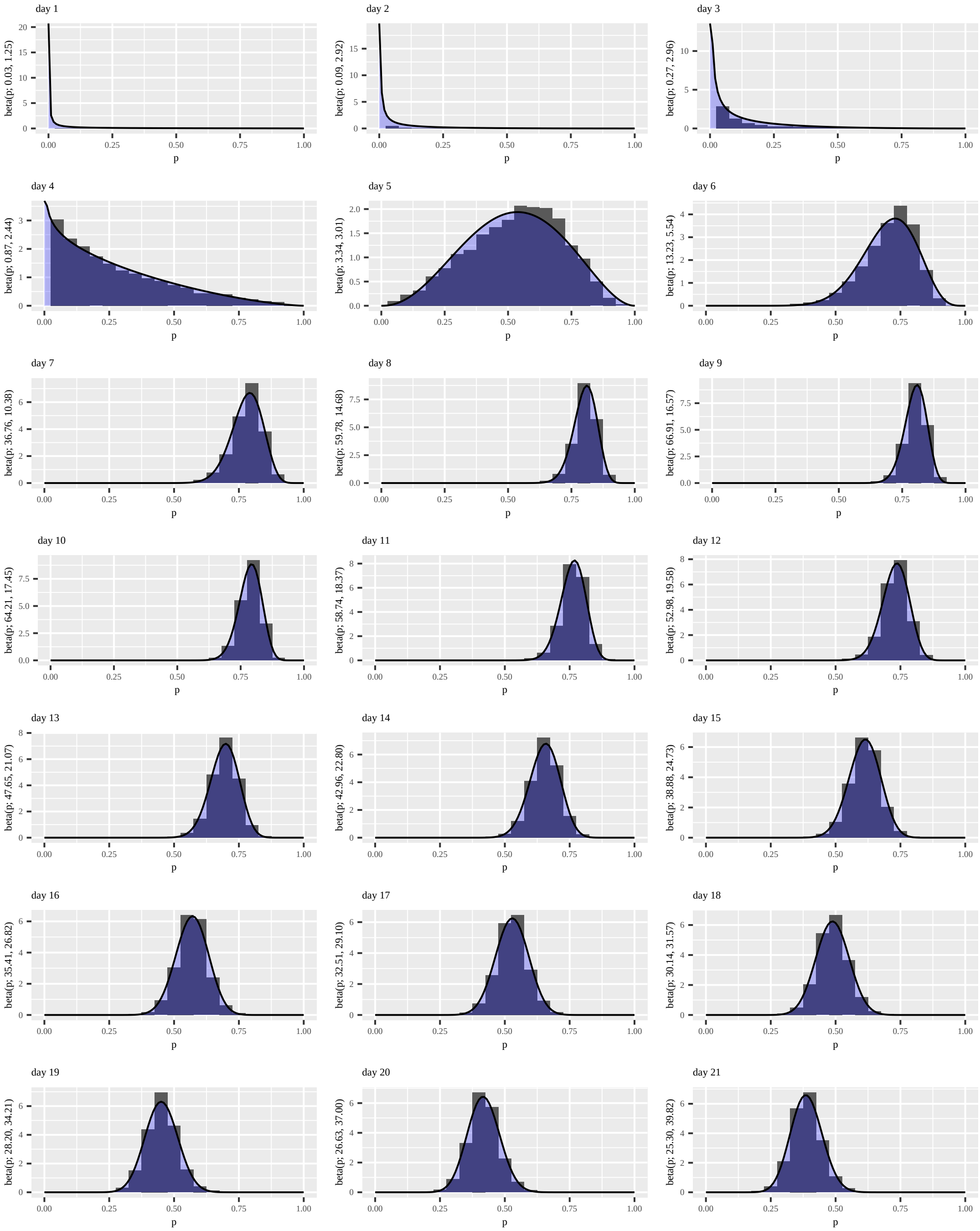


Figure 2: Daily sensitivity since day of infection of the PCR test assumed in this work.

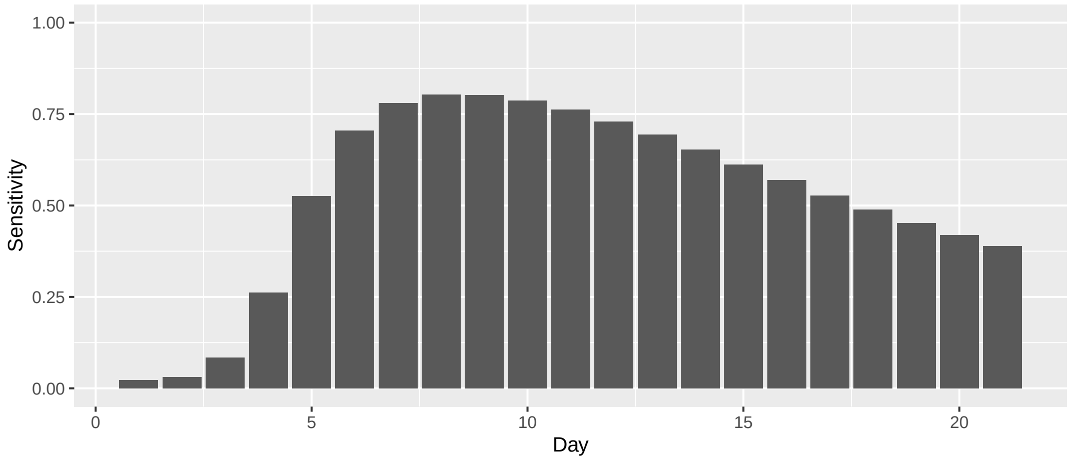


Figure 3: Daily mean sensitivity of the PCR test assumed in this work.

to the rooms within the office and one ACPH exchanged between the HVAC system and the external atmosphere. The air is distributed via ducts and returns to the HVAC via the ceiling plenum. Based on [19], this appears to be a standard air conditioning scenario for an office environment in Australia.

#### 4.2 Questacon

The museum is divided into public spaces and staff areas (see diagrams in Appendix B). Our analysis only considers the public spaces. The total floor space available to the public is about 5,000 $m^2$, with a corresponding volume of about 28,000 $m^3$, reflecting the high ceilings throughout the building.

The public space is centred around a large foyer, which contains entrance areas, ticketing and information desks, a shop, tables and seating for the adjacent café, and the museum shop. Other indoor public spaces, which are accessed from the foyer, are: the gallery block (detailed below); the café; and the Japan Theatre.

The core of the Questacon is a block of seven galleries arranged in a helical structure around an eighth cylindrical gallery ("The Drum"). A helical ramp on the outside of The Drum connects the galleries, which are numbered sequentially from top to bottom. The normal visitor path is to follow a long ramp from the foyer to the entrance to Gallery 1 (G1), the highest gallery, and thence down the ramp to subsequent galleries. The peripheral galleries are divided into three levels:

- Level 2: G1, Gallery 2 (G2) and Gallery 3 (G3), which have lateral links;
- Level 1: Gallery 4 (G4) and Gallery 5 (G5), which have a lateral link, and the mezzanine level of Gallery 7 (G7); and
- Ground Level: Gallery 6 (G6) and G7 and the central Gallery 8 (G8), all accessed from the Foyer.

Prior to COVID-19, from 2016 to 2019 the museum had between 487,691 and 524,263 visitors annually, with an average of about 1,400 visitors daily [13, 14, 15, 16]. Total visitor numbers for 2020 are not listed in the 2020 Annual Report, which notes considerable disruption due to COVID-19, including: prolonged closure to public access and other effects of the pandemic, such as public health restrictions, customer hesitancy and border closures [17]. Questacon had 11,803 and 8,746 visitors in June and July 2021 (respectively) [data provided by Questacon staff]. At this time the Australian Capital Territory (ACT) had not been significantly affected by the Delta-variant outbreak in NSW. These correspond to averages of 393 and 282 per day. Questacon also hosts school groups, but although they are included in the overall visitor numbers, we do not explicitly model school groups or other large groups. Only groups of up to 5 are modelled as a cohort.

During normal operations Questacon is open 9 AM to 5 PM every day of the year except Christmas Day. Periodically the Centre also hosts evening sessions including Questacon By Night.

The 2022 Questacon reopening plan, which runs to the end of June 2022, requires visitors to book for sessions commencing every hour from 9 AM onward. On most days the last session commences at 3 PM (giving 7 sessions per day), although this is extended to 7 PM on weekdays in February and March when school groups are planned (11 sessions per day). From 15 January up to 50 people will be allowed per session, corresponding to 350 for a normal day of operation, or up to 550 visitors during February and March. Questacon staff report the average visit is around 2 hours. The model assumes a return to normal operational capacity, which is 1,400 visitors per day and 9am to 5pm operation.

Staff are distributed within the public areas as follows:

- Front of house/Information desk: 4
- Café: 3
- Ticketing desk: 2
- Japan theatre: 1
- G1, G2, G3, G4, G5, G6 and G8: 1 per gallery
- Rover covering all galleries: 1
- Exhibition maintenance: 1 covering all galleries

In the current work we ignore the rover and the exhibition maintenance person.

G6 (Mini Q) and G7 (Science Sprouts) were not used during June/July 2021. The Questacon website indicates that currently Science Sprouts is closed and visitors to Mini Q are restricted. The current structure of our framework makes it difficult to represent the restrictions to Mini Q and we omit them. We further note that, when simulating a period, the activities and locations for all visitors are calculated, *then* these are aggregated to locations, meaning that at the time an agent 'makes the decision' to visit a room, the number of people in the room is not readily available and limits to room occupancy are not specifically modelled..

In our representation of the gallery complex we have divided the ramp around The Drum into seven segments connected to each of the seven peripheral galleries. All of these segments are connected to the central Drum (G8) These segments are used in the airflow model, but we do not include shedding or viral exposure in them as people spend minimal time on these segments while traversing between the galleries. The airflow across The Drum segments between the peripheral galleries and G8 is approximated, as is the airflow from the Foyer to G8.

The Japan Theatre and Blue Door Room (BDR) (a theatrette in G5) are self-contained with their own dedicated HVAC systems. With closed doors they have minimal air exchange with the Foyer or G5 (respectively). Each expels stale air directly to the outside, which is replaced via their HVAC systems. The 2022 Questacon reopening plan indicates the capacity of the Japan Theatre will be limited to 77 (it has 120 seats) and the BDR limited to 40. The main use of the BDR is by students, which we don't include here, and hence we omit it.

The café is open to the foyer. We have had some difficulty reconciling the café HVAC system and café areas and have omitted air exchange between the café and the Foyer.

G1, G2 and G3 have linking corridors with stairs (G1 to G2 and G2 to G3), as do G4 and G5 (G4 to G5). These enable visitor passage without having to return to The Drum ramp. Each is served by a dedicated HVAC unit, providing an "air wall" separating the two adjoining galleries. We were unable to determine the specifications for these units and so omit the links from our model.

Several small public spaces have been omitted in this model. Notably there are public toilets accessible through the foyer, in the Japan Theatre, on the ground and mezzanine level of G7, and off G3. We consider the additional risk of these facilities low as individual visits are relatively brief. One of us measured carbon dioxide levels in the Foyer toilets. The low concentrations suggested good ventilation despite the presence of a source in a confined space.

There are other rooms in the complex which we have ignored as they are unlikely to have a significant impact on our results under the well mixed room assumption. These rooms are relatively small, people do not tend to spend significant time in them, and they are well connected to the larger galleries. It is, however, possible that these rooms could present high risk areas for direct contact between individuals, which is not included in our model.

Some exhibits, such as the Earthquake Room are small spaces in themselves. They are noteworthy for having a staff member who talks to visitors in relative close proximity. We note that: Questacon applies the same density limits to these spaces as elsewhere; visitors spend relatively brief periods in these spaces; and these rooms are open to surrounding gallery, rather than closed; hence it is unlikely virus particles will concentrate in these spaces. In the current work we exclude these small exhibits, but again, they may present elevated direct contact risks.

The characteristics of the rooms of Questacon are shown in Table 4, and the flows between them are shown in Table 5 in Appendix B.

The HVAC system in the Questacon Galleries is quite sophisticated. In particular, it responds to Carbon Dioxide ($CO_2$) levels within each room. The default setting is 10% of the total throughput to be fresh air drawn from the outside. The same volume of air is expelled through a relief system. At a set point of 600 ppm it begins opening a damper allowing a greater proportion of fresh air into the circulation, with a limit of 100% of the air drawn from outside. For example G1, G2, G3, G6 and G7 each have a pair of Air Handling Units (AHUs) with a throughput of 10,800 $m^3/h$ of air, giving a total for each gallery of 21,600 $m^3/h$. Depending on $CO_2$ levels between 2,160 and 21,600 $m^3/h$ will be fresh air.

All air passing through the AHU is filtered before delivery into the gallery spaces. SARS-CoV-2 virions are thought to be concentrated in respiratory aerosols between 1 µm and 5 µm. The dense smoke from the early 2020 bushfires penetrated through the existing F5 filters (Minimum Efficiency Reporting Value (MERV) 8-9) installed in the Questacon HVAC systems, resulting in an unpleasant staff and visitor experience. Subsequently the filters were upgraded to F7 (MERV 13-14), the highest grade that can be fitted to the current HVAC systems. Bushfire smoke is dominated by PM2.5 particles (2.5 µm or less). These particles have the same size range as the respiratory aerosols transmitting COVID-19. MERV 8 filters capture only 20% of particles in the 1 to 3 µm range. MERV 13 filters capture 85% of such particles.

For the purposes of this report we assume each public space in Questacon receives 10% fresh air. The remaining 90% of the total output of each rooms HVAC system is drawn through a return duct within the room. Small inter-room flows are estimates that have been calculated as follows: a volume equal to 55% of the fresh air flow into G8 leaves the chamber via other galleries or the foyer; 10% each goes to the foyer, G7 (including the mezzanine level) and G6 (180 $m^3/h$). G1, G2, G3, G4 and G5 each receive 5% each (90 $m^3/h$). Flow from each peripheral gallery (G1 to G7 plus the G7 mezzanine) to G8 is assumed to be 10% of the total flow exiting the gallery. The remaining 90% is relief air venting directly to the outside. The return flow from the foyer to G8 is set equal and opposite at 180 $m^3/h$.

For example, G4 has a supply HVAC of 21,600 $m^3/h$. Of this 10% is fresh (2,160 $m^3/h$) and 90% is drawn from the room itself (19,440 $m^3/h$). An additional flow of 90 $m^3/h$ enters from the drum (G8) after flowing over the drum ramp outside G8. Hence the total flow entering G4 is 21,690 $m^3/h$, of which 10% (225 $m^3/h$) flows to G8 over the drum ramp and 90% (2,025 $m^3/h$) is exhausted directly to the exterior.

In total 990 $m^3/h$ leaves G8 via other galleries and the foyer, whilst 1,648 $m^3/h$ enters from other galleries and the foyer. Such a net inflow would implies the drum operates a a slight negative pressure.

The G6 logic was there no gallery above it (anticlockwise) on the drum ramp as the workshop and loading dock is on the other side. The drum ramp for G6 is defined as twice the area (& volume) of the drum ramps for the other galleries, except G7 which is double level. This was a fudge to approximately balance this drum ramp flow with the other segment. It's not necessary if visitors are assumed to spend no time on the drum ramp.

### 5 Air Filter Standards

| Australian Standard Ratings: AS1324.1-2001 & AS4260-1997 | | | European Ratings: EN779/EN1882 | U.S. MERV Ratings: ASHRAE 52.1 & 52.2 |
|---|---|---|---|---|
| Performance Rating | Average Arrestance No. 4 Test Dust | Average Efficiency No. 1 Test Dust | Performance Rating | Performance Rating |
| G1 G2 | < 65% 65%-80% | - | G1 G2 | MERV 1-4 |
| G3 | 80%-90% | - | G3 | MERV 5 |
| G4 | >90% | - | G4 | MERV 6-8 |
| F5 | - | 40%-60% | M5 | MERV 8-9 |
| F6 | - | 60%-80% | M6 | MERV 10-13 |
| F7 | - | 80%-90% | F7 | MERV 13-14 |
| F8 | - | 90%-95% | F8 | MERV 15 |
| F9 | - | >95% | F9 | MERV 16 |
| 95% DOP | - | - | H11 | - |
| Grade 1 | 99.97% 0.3µm | | H13 | - |
| Grade 2 | 99.99% 0.3µm | | - | - |
| Grade 3 | 99.999% 0.3µm | | H14 | - |
| Grade 4 | 99.999% 0.12µm | | U15 | - |

Table 1: The performance ratings given above are adapted from equivalent EU and MERV ratings and test classifications for filters as outlined in AS1324.1, EN779 and ASHRAE52. Adapted from [2].

### A Eveleigh

| Room | Use | Volume ($m^3$) | Room | Use | Volume ($m^3$) | Room | Use | Volume ($m^3$) |
|---|---|---|---|---|---|---|---|---|
| L4.37 | stair well | 242 | L5.24 | plant room | 52 | L5.52 | office | 38 |
| L4.63 | kitchen | 593 | L5.25 | stair well | 52 | L5.53 | office | 54 |
| L4.68b | corridor | 65 | L5.26a | meeting room | 280 | L5.56 | stair well | 32 |
| L4.L1 | lift | 11 | L5.26b | office | 62 | L5.57 | plant room | 51 |
| L4.L2 | lift | 11 | L5.27 | office | 35 | L5.58 | office | 32 |
| L4.L3 | lift | 11 | L5.28 | office | 53 | L5.59 | office | 41 |
| L5.01 | office | 52 | L5.29 | office | 36 | L5.63 | meeting room | 213 |
| L5.02 | office | 50 | L5.30 | office | 36 | L5.64 | office | 38 |
| L5.03 | office | 35 | L5.31 | foyer | 185 | L5.65 | office | 39 |
| L5.04 | office | 35 | L5.32 | reception | 32 | L5.66 | office | 38 |
| L5.05 | office | 35 | L5.33 | stair well | 274 | L5.67 | office | 54 |
| L5.06 | meeting room | 54 | L5.34 | alcove | 103 | L5.69 | open plan | 72 |
| L5.06a | alcove | 20 | L5.35 | plant room | 84 | L5.69a | open plan | 261 |
| L5.07 | office | 35 | L5.36 | office | 45 | L5.69b | corridor | 70 |
| L5.08 | office | 69 | L5.37 | meeting room | 55 | L5.69c | open plan | 292 |
| L5.09 | male toilet | 35 | L5.38 | office | 36 | L5.69d | open plan | 238 |
| L5.12 | stair well | 45 | L5.39 | meeting room | 35 | L5.69e | corridor | 105 |
| L5.13 | plant room | 13 | L5.40 | meeting room | 35 | L5.69g | corridor | 32 |
| L5.14 | male toilet | 34 | L5.41 | office | 117 | L5.69h | corridor | 72 |
| L5.16 | office | 38 | L5.43 | office | 35 | L5.70a | corridor | 100 |
| L5.17 | office | 38 | L5.44 | meeting room | 52 | L5.70b | corridor | 102 |
| L5.18 | office | 20 | L5.45 | male toilet | 37 | L5.70c | open plan | 302 |
| L5.21 | misc | 76 | L5.48 | stair well | 45 | L5.71a | corridor | 48 |
| L5.22 | office | 19 | L5.49 | plant room | 14 | L5.71b | open plan | 459 |
| L5.23 | office | 56 | L5.50 | male toilet | 35 | L5.72 | office | 37 |

Table 2: Characteristics of rooms used in the Eveleigh model. Note that none of these exchange air with the outside atmosphere directly; there is an also a single HVAC unit (not included).

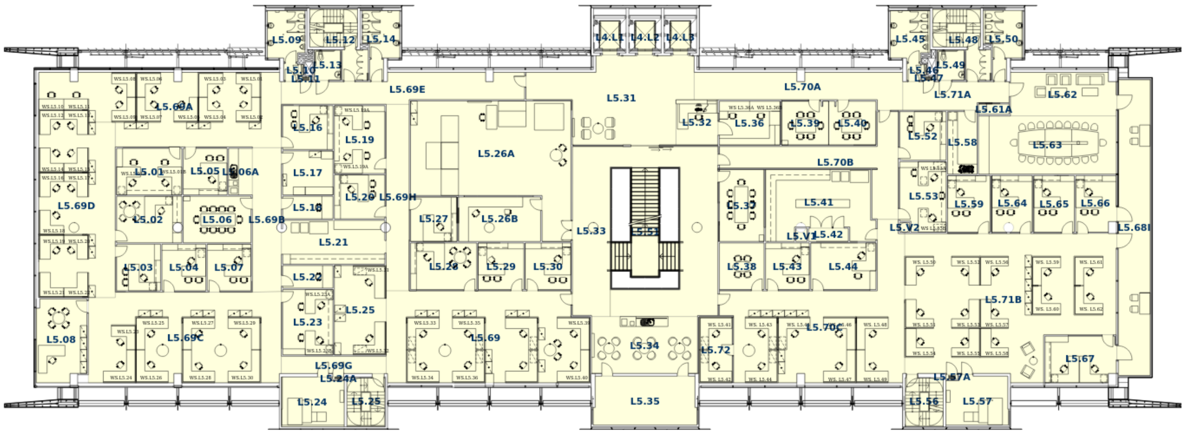


Figure 4: Floor plan of level 5 of the CSIRO office in Eveleigh.

| From | To | Rate ($m^3$/h) | From | To | Rate ($m^3$/h) | From | To | Rate ($m^3$/h) |
|---|---|---|---|---|---|---|---|---|
| L4.37 | L4.68b | 195 | L5.69h | L5.69 | 216 | L5.01 | L5.69a | 31 |
| L4.68b | L4.63 | 195 | L5.69h | L5.27 | 77 | L5.05 | L5.69a | 94 |
| L5.06 | L5.33 | 161 | L5.70b | L5.56 | 3 | L5.16 | L5.69a | 82 |
| L5.08 | L5.69c | 114 | L5.70b | L5.57 | 5 | L5.06 | L5.69b | 145 |
| L5.26a | L5.26b | 168 | L5.70b | L5.70a | 300 | L5.06a | L5.69b | 61 |
| L5.26a | L5.33 | 596 | L5.70b | L4.L1 | 13 | L5.21 | L5.69b | 210 |
| L5.31 | L5.32 | 97 | L5.70b | L5.52 | 43 | L5.22 | L5.69b | 41 |
| L5.31 | L5.33 | 93 | L5.70b | L5.53 | 32 | L5.18 | L5.69b | 22 |
| L5.31 | L5.39 | 58 | L5.70c | L5.70b | 305 | L5.17 | L5.69b | 23 |
| L5.31 | L5.40 | 95 | L5.70c | L5.72 | 42 | L5.69d | L5.69c | 713 |
| L5.31 | L5.70a | 300 | L5.70c | L5.41 | 192 | L5.69b | L5.69c | 210 |
| L5.31 | L5.71a | 144 | L5.70c | L5.44 | 113 | L5.69g | L5.69c | 96 |
| L5.31 | L4.L1 | 6 | L5.70c | L5.43 | 94 | L5.04 | L5.69c | 58 |
| L5.31 | L4.L2 | 6 | L5.70c | L5.38 | 21 | L5.07 | L5.69c | 21 |
| L5.31 | L4.L3 | 6 | L5.71a | L5.45 | 18 | L5.23 | L5.69c | 92 |
| L5.32 | L5.36 | 87 | L5.71a | L5.49 | 1 | L5.69a | L5.69d | 713 |
| L5.33 | L5.34 | 278 | L5.71a | L5.48 | 4 | L5.02 | L5.69d | 136 |
| L5.33 | L5.70b | 305 | L5.71a | L5.50 | 18 | L5.03 | L5.69d | 21 |
| L5.33 | L5.37 | 119 | L5.71a | L5.58 | 69 | L5.09 | L5.69e | 17 |
| L5.33 | L4.37 | 725 | L5.71a | L5.63 | 130 | L5.13 | L5.69e | 1 |
| L5.34 | L5.35 | 8 | L5.71b | L5.56 | 3 | L5.12 | L5.69e | 4 |
| L5.69 | L5.28 | 59 | L5.71b | L5.57 | 5 | L5.14 | L5.69e | 17 |
| L5.69 | L5.29 | 22 | L5.71b | L5.67 | 89 | L5.69h | L5.69e | 216 |
| L5.69 | L5.30 | 22 | L5.71b | L5.59 | 25 | L5.31 | L5.69e | 53 |
| L5.69 | L5.33 | 119 | L5.71b | L5.64 | 82 | L5.24 | L5.69g | 3 |
| L5.69a | L5.69b | 210 | L5.71b | L5.65 | 23 | L5.25 | L5.69g | 3 |
| L5.69a | L5.69e | 316 | L5.71b | L5.66 | 43 | L5.69 | L5.69g | 96 |
| L5.69a | L5.01 | 31 | L4.68b | L4.37 | 195 | L5.69 | L5.69h | 216 |
| L5.69a | L5.05 | 94 | L4.63 | L4.68b | 195 | L5.27 | L5.69h | 77 |
| L5.69a | L5.16 | 82 | L5.33 | L5.06 | 161 | L5.56 | L5.70b | 3 |
| L5.69b | L5.06 | 145 | L5.69c | L5.08 | 114 | L5.57 | L5.70b | 5 |
| L5.69b | L5.06a | 61 | L5.26b | L5.26a | 168 | L5.70a | L5.70b | 300 |
| L5.69b | L5.21 | 210 | L5.33 | L5.26a | 596 | L4.L1 | L5.70b | 13 |
| L5.69b | L5.22 | 41 | L5.32 | L5.31 | 97 | L5.52 | L5.70b | 43 |
| L5.69b | L5.18 | 22 | L5.33 | L5.31 | 93 | L5.53 | L5.70b | 32 |
| L5.69b | L5.17 | 23 | L5.39 | L5.31 | 58 | L5.70b | L5.70c | 305 |
| L5.69c | L5.69d | 713 | L5.40 | L5.31 | 95 | L5.72 | L5.70c | 42 |
| L5.69c | L5.69b | 210 | L5.70a | L5.31 | 300 | L5.41 | L5.70c | 192 |
| L5.69c | L5.69g | 96 | L5.71a | L5.31 | 144 | L5.44 | L5.70c | 113 |
| L5.69c | L5.04 | 58 | L4.L1 | L5.31 | 6 | L5.43 | L5.70c | 94 |
| L5.69c | L5.07 | 21 | L4.L2 | L5.31 | 6 | L5.38 | L5.70c | 21 |
| L5.69c | L5.23 | 92 | L4.L3 | L5.31 | 6 | L5.45 | L5.71a | 18 |
| L5.69d | L5.69a | 713 | L5.36 | L5.32 | 87 | L5.49 | L5.71a | 1 |
| L5.69d | L5.02 | 136 | L5.34 | L5.33 | 278 | L5.48 | L5.71a | 4 |
| L5.69d | L5.03 | 21 | L5.70b | L5.33 | 305 | L5.50 | L5.71a | 18 |
| L5.69e | L5.09 | 17 | L5.37 | L5.33 | 119 | L5.58 | L5.71a | 69 |
| L5.69e | L5.13 | 1 | L4.37 | L5.33 | 725 | L5.63 | L5.71a | 130 |
| L5.69e | L5.12 | 4 | L5.35 | L5.34 | 8 | L5.56 | L5.71b | 3 |
| L5.69e | L5.14 | 17 | L5.28 | L5.69 | 59 | L5.57 | L5.71b | 5 |
| L5.69e | L5.69h | 216 | L5.29 | L5.69 | 22 | L5.67 | L5.71b | 89 |
| L5.69e | L5.31 | 53 | L5.30 | L5.69 | 22 | L5.59 | L5.71b | 25 |
| L5.69g | L5.24 | 3 | L5.33 | L5.69 | 119 | L5.64 | L5.71b | 82 |
| L5.69g | L5.25 | 3 | L5.69b | L5.69a | 210 | L5.65 | L5.71b | 23 |
| L5.69g | L5.69 | 96 | L5.69e | L5.69a | 316 | L5.66 | L5.71b | 43 |

Table 3: Inter-room air exchange used in the Eveleigh model.

### B Questacon

| Box | Volume ($m^3$) | External Venting out ($m^3/h$) | External Venting in ($m^3/h$) |
|---|---|---|---|
| G1 | 2695 | 2025 | 0 |
| G2 | 3185 | 2025 | 0 |
| G3 | 3675 | 2025 | 0 |
| G4 | 1752 | 2025 | 0 |
| G5 | 1197 | 904 | 0 |
| G5_BDR | 437 | 648 | 0 |
| G6 | 1307 | 2106 | 0 |
| G7 | 2415 | 2106 | 0 |
| G8 | 3296 | 2458 | 0 |
| cafe | 472 | 1886 | 0 |
| dr_G1 | 118 | 0 | 0 |
| dr_G2 | 125 | 0 | 0 |
| dr_G3 | 143 | 0 | 0 |
| dr_G4 | 136 | 0 | 0 |
| dr_G5 | 136 | 0 | 0 |
| dr_G6 | 271 | 0 | 0 |
| dr_G7 | 100 | 0 | 0 |
| dr_G7M | 136 | 0 | 0 |
| foyer | 5880 | 4608 | 0 |
| h_G1 | 1 | 0 | 2160 |
| h_G2 | 1 | 0 | 2160 |
| h_G3 | 1 | 0 | 2160 |
| h_G4 | 1 | 0 | 2160 |
| h_G5 | 1 | 0 | 914 |
| h_G5_BDR | 1 | 0 | 648 |
| h_G6 | 1 | 0 | 2160 |
| h_G7 | 1 | 0 | 2160 |
| h_G8 | 1 | 0 | 1800 |
| h_cafe | 1 | 0 | 1886 |
| h_foyer | 1 | 0 | 4608 |
| h_japan_thtr | 1 | 0 | 864 |
| japan_thtr | 1134 | 864 | 0 |

Table 4: Characteristics of boxes used in the Questacon model. The column "External Venting out" contains the volumes of air vented from the box to the external atmosphere, and "External Venting in" contains the volumes of air pumped into the boxes from the external Atmosphere.

| From | To | Rate ($m^3/h$) | From | To | Rate ($m^3/h$) |
|---|---|---|---|---|---|
| Gallery 1 | Gallery 1 Drum | 225 | Gallery 2 Drum | Gallery 2 | 90 |
| Gallery 1 | Gallery 1 HVAC | 19440 | Gallery 2 Drum | Gallery 8 | 225 |
| Gallery 2 | Gallery 2 Drum | 225 | Gallery 3 Drum | Gallery 3 | 90 |
| Gallery 2 | Gallery 2 HVAC | 19440 | Gallery 3 Drum | Gallery 8 | 225 |
| Gallery 3 | Gallery 3 Drum | 225 | Gallery 4 Drum | Gallery 4 | 90 |
| Gallery 3 | Gallery 3 HVAC | 19440 | Gallery 4 Drum | Gallery 8 | 225 |
| Gallery 4 | Gallery 4 Drum | 225 | Gallery 5 Drum | Gallery 5 | 90 |
| Gallery 4 | Gallery 4 HVAC | 19440 | Gallery 5 Drum | Gallery 8 | 100 |
| Gallery 5 | Gallery 5 Drum | 100 | Gallery 6 Drum | Gallery 6 | 180 |
| Gallery 5 | Gallery 5 HVAC | 8230 | Gallery 6 Drum | Gallery 8 | 234 |
| Blue Door Room | Blue Door Room HVAC | 5832 | Gallery 7 Drum | Gallery 7 | 90 |
| Gallery 6 | Gallery 6 Drum | 234 | Gallery 7 Drum | Gallery 8 | 117 |
| Gallery 6 | Gallery 6 HVAC | 19440 | Gallery 7 Mezanine Drum | Gallery 7 | 90 |
| Gallery 7 | Gallery 7 Drum | 117 | Gallery 7 Mezanine Drum | Gallery 8 | 117 |
| Gallery 7 | Gallery 7 Mezanine Drum | 117 | Foyer | Gallery 8 | 180 |
| Gallery 7 | Gallery 7 HVAC | 19440 | Foyer | Foyer HVAC | 41472 |
| Gallery 8 | Gallery 1 Drum | 90 | Gallery 1 HVAC | Gallery 1 | 21600 |
| Gallery 8 | Gallery 2 Drum | 90 | Gallery 2 HVAC | Gallery 2 | 21600 |
| Gallery 8 | Gallery 3 Drum | 90 | Gallery 3 HVAC | Gallery 3 | 21600 |
| Gallery 8 | Gallery 4 Drum | 90 | Gallery 4 HVAC | Gallery 4 | 21600 |
| Gallery 8 | Gallery 5 Drum | 90 | Gallery 5 HVAC | Gallery 5 | 9144 |
| Gallery 8 | Gallery 6 Drum | 180 | Blue Door Room HVAC | Blue Door Room | 6480 |
| Gallery 8 | Gallery 7 Drum | 90 | Gallery 6 HVAC | Gallery 6 | 21600 |
| Gallery 8 | Gallery 7 Mezanine Drum | 90 | Gallery 7 HVAC | Gallery 7 | 21600 |
| Gallery 8 | Foyer | 180 | Gallery 8 HVAC | Gallery 8 | 18000 |
| Gallery 8 | Gallery 8 HVAC | 16200 | Cafe HVAC | Cafe | 18864 |
| Cafe | Cafe HVAC | 16978 | Foyer HVAC | Foyer | 46080 |
| Gallery 1 Drum | Gallery 1 | 90 | Japan Theatre HVAC | Japan Theatre | 8640 |
| Gallery 1 Drum | Gallery 8 | 225 | Japan Theatre | Japan Theatre HVAC | 7776 |

Table 5: Inter-room air exchange used in the Questacon model.

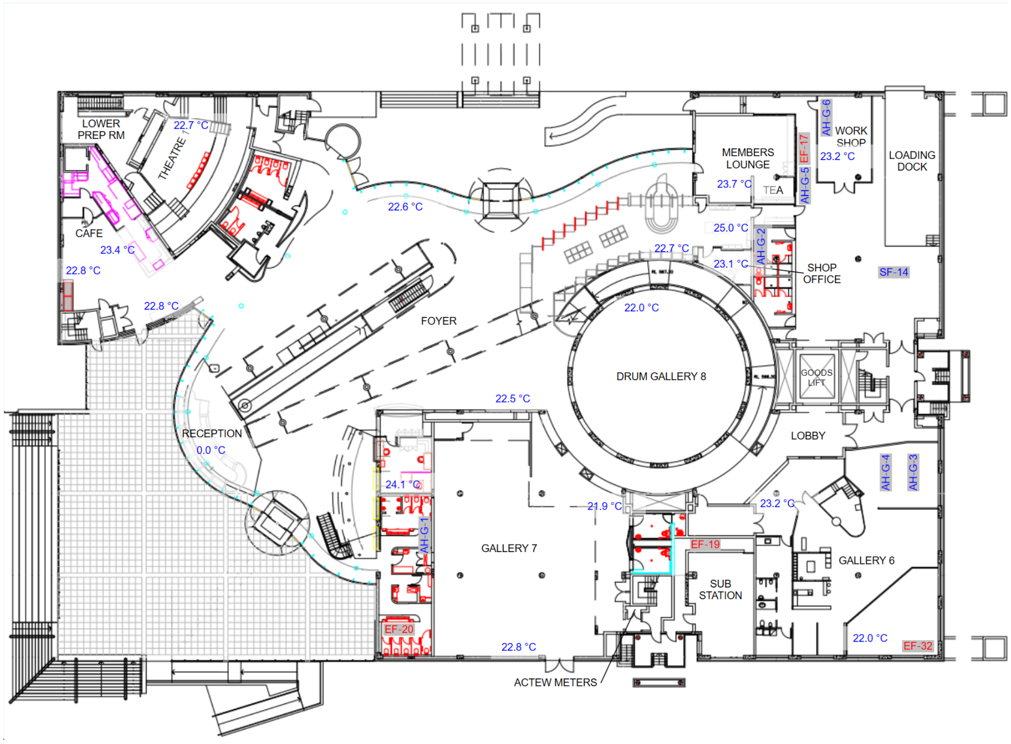


Figure 5: Floor plan of the ground floor of Questacon.

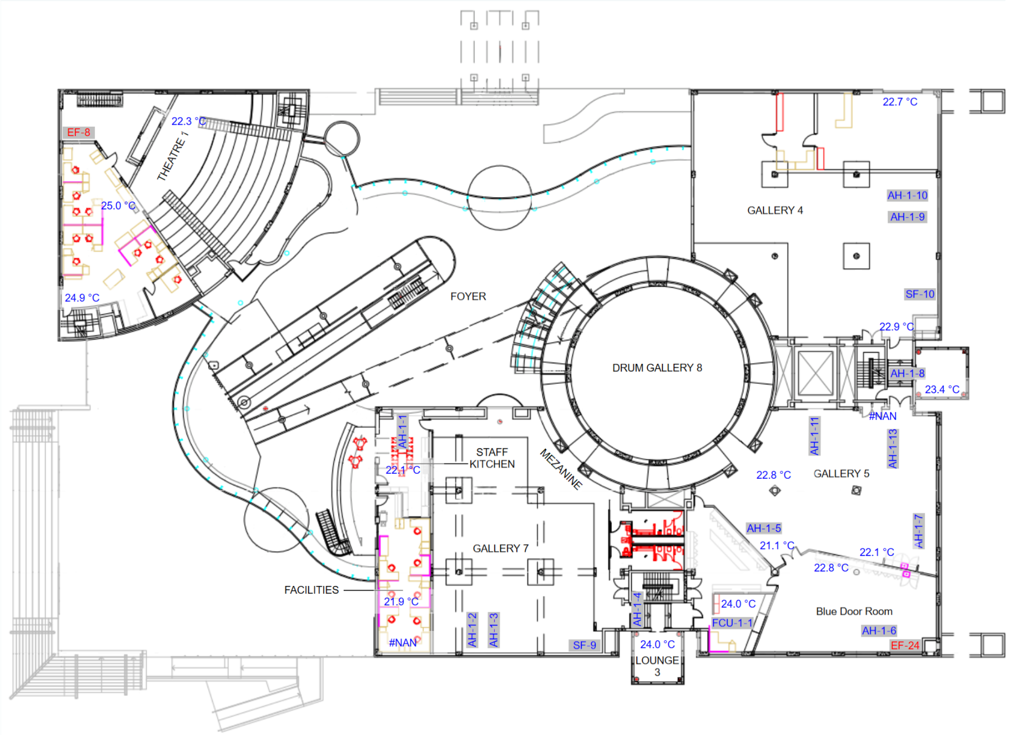


Figure 6: Floor plan of level 1 of Questacon.

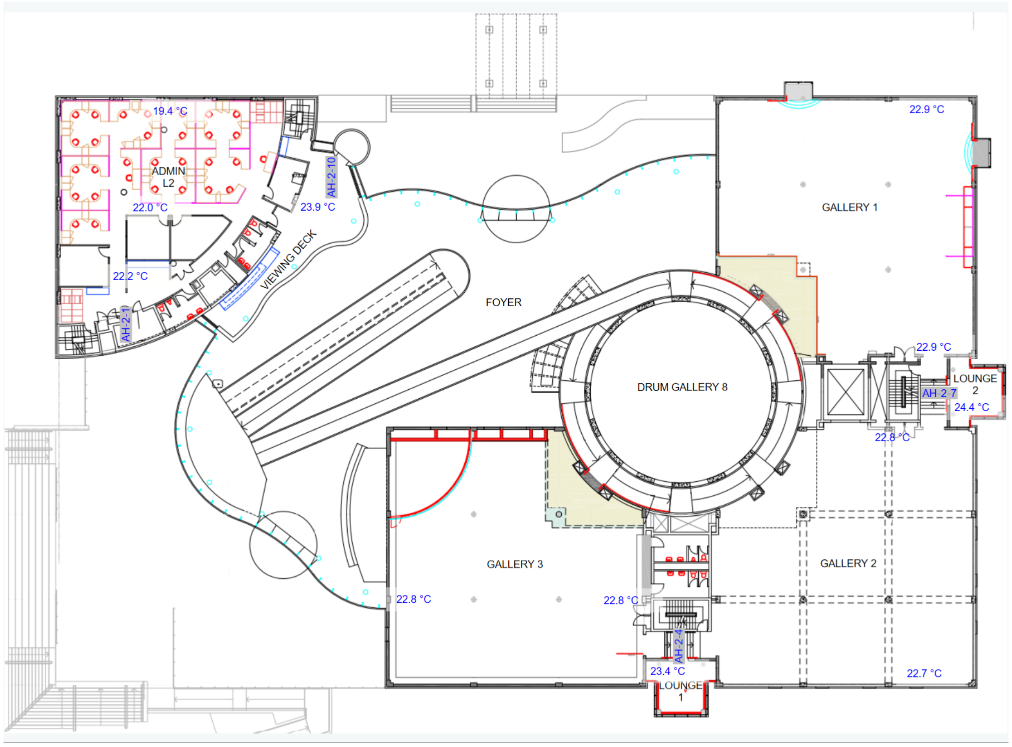


Figure 7: Floor plan of level 2 of Questacon.
